# Catatonic Features That May Be Mistaken for Worsening Autism Show Treatment-Associated Improvement: Implications for Later Regression

**DOI:** 10.64898/2026.09.14.26363007

**Authors:** Joshua Ryan Smith, Maria Bonnee, Sarah Marler, Seri Lim, D. Catherine Fuchs, Rafael Tamargo, Isaac Baldwin, Christopher Maley, Ashley VanHaverbeck, Courtney Hamilton, Timothy Adegoke, Haozheng Xu, Jinyuan Liu, Zachary J. Williams, Jo Ellen Wilson, James Luccarelli

## Abstract

Catatonia in autistic individuals may resemble worsening autism yet represent a potentially treatable departure from baseline functioning. We compared item-level catatonic signs in autistic and non-autistic patients and evaluated treatment-associated change during electroconvulsive therapy (ECT). This single-center observational cohort included patients with catatonia who received ECT from May 2022 through April 2026. The 23-item Bush-Francis Catatonia Rating Scale (BFCRS) was assessed longitudinally. Pretreatment BFCRS data were available for 104 patients, including 41 autistic and 63 non-autistic patients, with 993 near-complete repeated assessments available. Pretreatment item presence was modeled using a three-level clinical-group variable with adjustment for age, biologic sex, and calendar year. Compared with non-autistic patients, patients with autism and intellectual disability had higher adjusted odds of eight activated, repetitive, or behaviorally dysregulated signs and lower odds of immobility/stupor. Among 96 patients with paired first and last eligible BFCRS assessments, multiple items improved in both cohorts. Generalized estimating equations demonstrated longitudinal declines in total BFCRS scores and multiple psychomotor-domain scores, with no clinical-group-by-time interaction surviving false-discovery rate correction in unrestricted or 30-, 60-, 90-, and 180-day models. Autism-specific Kanner analyses identified significant fixed-endpoint improvement in seven severity items after false-discovery rate correction, six of which were reproduced longitudinally. Catatonia in autistic patients with intellectual disability was characterized by a phenotype that may resemble worsening autism during later regression, and many constituent signs demonstrated treatment-associated improvement during ECT.

**Lay Summary:** Some autistic individuals experience later behavioral, motor, or functional regression, which may have potentially treatable causes such as catatonia. In this study, patients with autism and intellectual disability were more likely to show activated, repetitive, and behaviorally dysregulated catatonic features than non-autistic patients, and these features improved during treatment. Catatonia should be considered when an autistic person develops new or substantially worsening repetitive behavior, impulsivity, aggression, communication loss, reduced self-care, mobility changes, food refusal, or other departures from prior functioning.

## Introduction

Catatonia is an increasingly recognized, high-morbidity psychomotor syndrome that occurs across the life cycle and disproportionately affects autistic individuals and those with genetic syndromes (1–10). In 2000, Wing and Shah conducted the seminal study demonstrating catatonia’s high prevalence in autism (11). If unrecognized and untreated, catatonia may progress to malignant catatonia, which is associated with autonomic instability and substantial morbidity and mortality (7,12). Early recognition and treatment, including pharmacotherapy and electroconvulsive therapy (ECT), are therefore imperative (1,3,7,13–16).

For autistic individuals, detecting catatonia is challenging because of symptom overlap with longstanding autism characteristics and difficulty identifying departures from prior functioning (8,17). A recent meta-analysis found that 20.2% of autistic individuals had features of catatonia, with 10.4% meeting full criteria (18,19). Common features of catatonia in autism include new-onset speech impairment, negativism, hyperactivity, recurrent self-injury, and aggression, further complicating diagnosis (18,20–22). Regression has traditionally been discussed in relation to autism onset during early childhood, typically involving loss or decline in language or social communication skills. A systematic review and meta-analysis estimated that approximately 30% of autistic children experience regression, with average onset during the second year of life, although prevalence varies substantially according to definition and measurement method (23,24). Later regression in an already diagnosed autistic individual presents a different clinical challenge. Loss of communication, self-care, mobility, continence, food intake, behavioral regulation, or previously acquired skills may result from psychiatric, neurologic, genetic, medical, environmental, or behavioral causes, including catatonia (17,25). Identifying whether observed behavior represents a meaningful departure from prior functioning is therefore central to distinguishing catatonia from longstanding autism-related features.

Although prominent activated features of catatonia in autism have been described, no study has systematically compared item-level catatonic signs in autistic and non-autistic patients using a standardized instrument such as the Bush-Francis Catatonia Rating Scale (BFCRS) (26). In a companion analysis of this cohort, autistic patients with catatonia received more ECT sessions over longer treatment courses than non-autistic patients (27). The present study aimed to identify pretreatment BFCRS features associated with autism and intellectual disability and determine whether these features changed during ECT. We hypothesized that increased psychomotor signs and selected repetitive or behaviorally dysregulated signs would be more prominent in patients with autism and intellectual disability, whereas classically hypokinetic signs would be more prominent in non-autistic patients. We further hypothesized that catatonic signs resembling baseline autism characteristics or worsening autism, but representing a departure from prior functioning, would demonstrate treatment-associated improvement. Such improvement would support recognition of catatonia as a potentially treatable contributor to later regression. Autism-specific analyses using the Kanner Catatonia Rating Scale (KCRS), comprising the Kanner Catatonia Severity scale (KCS) and Kanner Catatonia Examination (KCE), provided complementary assessments of change (28).

## Methods

### Study Population

Following *Strengthening the Reporting of Observational Studies in Epidemiology* (STROBE) guidelines (29), we conducted an observational cohort study of patients with catatonia who received ECT at Vanderbilt University Medical Center (VUMC). Data were managed with Research Electronic Data Capture (REDCap) tools hosted at VUMC (30,31). Patient information was collected longitudinally from May 11, 2022, to April 30, 2026.

For multivariable analyses, patients were classified as non-autism, autism without intellectual disability, or autism with intellectual disability. Autism was diagnosed or confirmed by the treating psychiatrist using Diagnostic and Statistical Manual of Mental Disorders, Fifth Edition (DSM-5) criteria; intellectual disability was defined by clinician diagnosis (32).

Patients were included in the present phenotype analysis if they had a diagnosis of catatonia, received at least two ECT treatments at an interval of 7 days or less, and had an evaluable pretreatment BFCRS assessment. Of the 110 patients included in the companion treatment-course cohort, 104 had an evaluable pretreatment near-complete BFCRS assessment and comprised the present phenotype sample; 96 of these patients had at least two eligible near-complete BFCRS assessments and contributed to paired and longitudinal analyses. In both cohorts, catatonia was diagnosed using DSM-5 criteria and assessed longitudinally using the BFCRS (26). In the autism cohort, catatonia was additionally assessed using the KCRS, comprising the KCS and KCE (28). Other psychiatric conditions that could produce overlapping presentations, including anxiety, psychotic, and mood disorders, were evaluated using an unstructured clinical psychiatric interview based on DSM-5 criteria conducted by a board-certified psychiatrist.

### ECT Treatment

Informed consent for ECT was obtained for all patients in accordance with Tennessee state law. The research study was reviewed by the VUMC Institutional Review Board (#242126) and granted a waiver of informed consent, as data were collected as part of routine clinical care. This study is a continuation of our previous work with adults and children with autism and intellectual disability (14,15,33), and pediatric catatonia more broadly (2,34–36). These item-level and longitudinal analyses have not been published previously.

ECT treatments were performed using a Thymatron IV instrument (Somatics, LLC). The initial stimulus dose was determined using the half-age method (37) and was subsequently increased as needed to achieve an adequate seizure. All treatments were performed under general anesthesia with neuromuscular blockade.

### Measures

The BFCRS and KCRS were recorded longitudinally during routine care by raters who were not blinded to diagnosis or treatment course (26,28). Signs were interpreted relative to typical pre-catatonia functioning, using caregiver input when available. Later behavioral, motor, or functional regression informed interpretation of change from baseline but was not analyzed as a study variable because its timing, tempo, affected domains, and relationship to treatment were not recorded systematically. Here, regression refers to clinical loss of functioning, not statistical regression modeling. “Pretreatment assessment” denotes the first eligible rating; “pre-catatonia functioning” denotes typical functioning before catatonia onset or worsening.

The BFCRS includes a 14-item screening instrument and a 23-item severity scale (26). Assessments with at least 20 scored items were considered near-complete, and total scores were prorated to 23 items. The first near-complete assessment was designated pretreatment. Item-level analyses used available-case denominators. The KCRS comprises the 18-item KCS and 12-item KCE (28,38). Eligible assessments required at least 15 KCS or 10 KCE items, and totals were prorated to the full scale.

BFCRS items were assigned to increased, abnormal, and decreased psychomotor-behavior domains according to the checklist presented by Walther and colleagues (39). The increased domain comprised excitement, impulsivity, and combativeness. The abnormal domain comprised posturing/catalepsy, grimacing, echopraxia/echolalia, stereotypy, mannerisms, verbigeration, automatic obedience, mitgehen, gegenhalten, grasp reflex, perseveration, rigidity, and waxy flexibility. The decreased domain comprised immobility/stupor, mutism, staring, negativism, withdrawal, and ambitendency. Autonomic abnormality was retained separately because it is not a psychomotor sign. These externally defined psychomotor-behavior domains were used to organize descriptive and longitudinal findings and were not interpreted as independently validated catatonia subtypes. For paired analyses, first and last occasions were selected at the scale level before item testing; item-specific denominators could vary because of missing endpoint values.

### Data Collection

Characteristics included age, biologic sex, intellectual disability, calendar year of ECT initiation, and consultation setting. Other clinical and treatment details are reported in the companion analysis (27). Because equivalent inpatient pathways were not readily available to autistic patients with intellectual disability, consultation setting was evaluated in structural-access sensitivity analyses rather than included in the primary adjustment set.

### Statistical Analyses

Statistical analyses were performed using Python version 3.12.4 (Python Software Foundation). All tests were two-sided, and statistical significance was defined as p<0.05. Benjamini-Hochberg false-discovery rate (FDR) correction was applied within each prespecified family of item-level tests, principal group-specific longitudinal slope tests, or longitudinal interaction tests, with q<0.05 considered statistically significant.

### Use of generative artificial intelligence

VUMC’s Microsoft 365 Copilot was used under author supervision to support statistical-code development and review, figure-code preparation, consistency checks, and manuscript editing. The authors reviewed and executed all code, verified reported outputs against the analytic data, critically revised all AI-assisted material, and remain responsible for the study methods, interpretation, originality, privacy compliance, and final content.

### Baseline BFCRS phenotype comparisons

For each of the 23 standard BFCRS items, symptom presence was defined as a score greater than zero. Unadjusted descriptive comparisons between the binary autism and non-autism cohorts were performed using two-sided Fisher exact tests. Benjamini-Hochberg FDR correction was applied across the 23 prevalence tests.

Primary pretreatment analyses used item-level multivariable binomial generalized linear models with symptom presence as the dependent variable, three-level clinical group as the primary predictor, and age at consultation, biologic sex, and calendar year of ECT initiation as covariates. Non-autism was the reference group. The primary contrasts compared autism with intellectual disability and autism without intellectual disability separately with non-autism. FDR correction was applied across the 46 prespecified subgroup contrasts, comprising two clinical-group contrasts for each of the 23 BFCRS items.

The autism-without-intellectual-disability subgroup contained five patients. Estimates for this subgroup were therefore considered exploratory regardless of statistical significance, and sparse or imprecise estimates were not interpreted as evidence of absence or protection. Consultation setting was added to sensitivity models to evaluate attenuation, preservation of effect direction, and dependence on referral context. A sensitivity analysis modeled age at consultation using a restricted cubic spline with three degrees of freedom while retaining the original 46-contrast FDR family.

### BFCRS treatment response analyses

Patients with at least two near-complete BFCRS assessments were included in fixed-endpoint analyses (N=96; 39 autistic and 57 non-autistic patients). The first and last near-complete assessment occasions were selected at the scale level before item-level analysis. Change was defined as the first assessment score minus the last assessment score, such that positive values represented improvement.

First and last scores were compared separately within the autism and non-autism cohorts using two-sided paired Wilcoxon signed-rank tests with Pratt handling of zero-difference ties (40). Benjamini-Hochberg FDR correction was applied separately across the 23 BFCRS item tests in each cohort. Rank-biserial correlations were calculated as paired nonparametric effect sizes, with positive values indicating lower scores at the final assessment.

These analyses evaluated within-cohort change and were not interpreted as estimates of ECT efficacy because there was no untreated comparison group and concurrent clinical interventions were not standardized.

### BFCRS secondary longitudinal analyses

Primary unrestricted longitudinal analyses used all eligible near-complete BFCRS assessments across each patient’s entire observed follow-up to estimate change over time while accounting for repeated observations and unequal assessment timing. Time was defined as days since the first eligible BFCRS assessment and transformed as ln(days+1) to accommodate denser observations early in treatment and longer intervals during maintenance ECT.

BFCRS items were organized into the increased, abnormal, and decreased psychomotor-behavior domains specified above according to the checklist presented by Walther and colleagues (39). Multi-item domain scores were calculated as mean item scores when at least 80% of constituent items were observed.

Population-averaged associations were estimated using generalized estimating equations (GEE) with an identity link, patient-level clustering, an exchangeable working-correlation structure, and robust standard errors. Model predictors included ln(days+1), three-level clinical group, the clinical-group-by-time interaction, age at consultation, biologic sex, and calendar year of ECT initiation. Non-autism was the reference group. Consultation setting was added in structural-access sensitivity models. Regression coefficients represent change per one-unit increase in ln(days+1).

Common-window sensitivity analyses repeated these models using only assessments within 30, 60, 90, and 180 days of each patient’s first eligible assessment. Additional unrestricted sensitivity analyses used an independence working-correlation structure or were restricted to complete assessments with all 23 BFCRS items observed and no score proration. Patients were required to have at least two eligible assessments in each analysis. For the unrestricted primary analysis, complete-assessment sensitivity analysis, independence working-correlation sensitivity analysis, and each common follow-up window, FDR correction was applied separately to the 10 principal group-specific slope tests, comprising five outcomes in the autism-with-intellectual-disability and non-autism groups. FDR correction was also applied to separate 10-test clinical-group-by-time interaction families for the unrestricted primary analysis, consultation-setting sensitivity analysis, complete-assessment sensitivity analysis, independence working-correlation sensitivity analysis, and each common follow-up window. Autism-without-intellectual-disability estimates remained exploratory.

### Autism-specific Kanner Catatonia Rating Scale Analysis

Kanner analyses were restricted to autistic patients and evaluated autism-specific treatment-associated change.

For KCS item-level analyses, autistic patients with at least two eligible KCS assessments were included. First and last eligible KCS assessment occasions were selected at the scale level, and graded item scores were compared using two-sided paired Wilcoxon signed-rank tests with Pratt handling of zero-difference ties (40). Rank-biserial correlations were calculated as effect sizes. FDR correction was applied across the 18 KCS item tests.

For KCE item-level analyses, binary examination-sign status at fixed first and last eligible assessment occasions was compared using two-sided exact McNemar tests. Resolved, newly emergent, persistent, and absent-at-both-assessments counts were summarized. FDR correction was applied across the 12 KCE item tests.

KCS longitudinal sensitivity analyses used generalized estimating equations (GEE) with patient-level clustering, an exchangeable working-correlation structure, robust standard errors, and adjustment for age, biologic sex, and calendar year of ECT initiation. Time was represented as the natural logarithm of one plus days since the first eligible Kanner assessment.

Post hoc longitudinal KCE models used binomial GEE and were fitted only for items with at least 10 positive observations, at least 10 negative observations, and at least five patients with within-patient outcome variation. FDR correction was applied across eligible, successfully fitted items.

## Results

### Relevant Demographic and Therapeutic Data

The pretreatment sample included 63 non-autistic and 41 autistic patients, including 36 autistic patients with intellectual disability and 5 without intellectual disability (Table 1). Additional clinical and treatment characteristics are reported in the companion analysis (27).

**Table 1:**
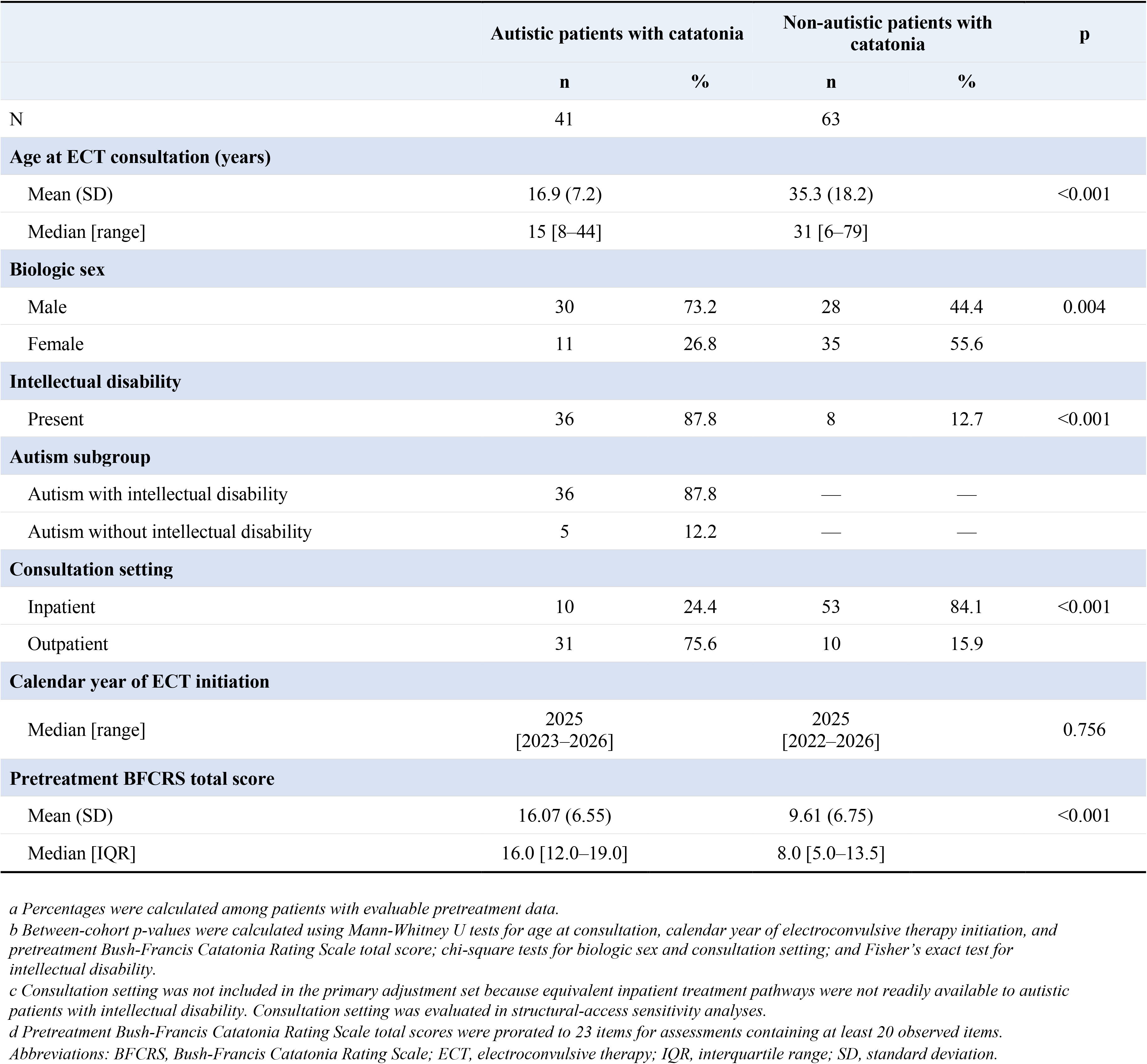
Pretreatment Sample Characteristics.

|  | Autistic patients with catatonia |  | Non-autistic patients with catatonia |  | p |
| --- | --- | --- | --- | --- | --- |
|  | n | % | n | % |  |
| N | 41 |  | 63 |  |  |
| Age at ECT consultation (years) |  |  |  |  |  |
| Mean (SD) | 16.9 (7.2) |  | 35.3 (18.2) |  | <0.001 |
| Median [range] | 15 [8–44] |  | 31 [6–79] |  |  |
| Biologic sex |  |  |  |  |  |
| Male | 30 | 73.2 | 28 | 44.4 | 0.004 |
| Female | 11 | 26.8 | 35 | 55.6 |  |
| Intellectual disability |  |  |  |  |  |
| Present | 36 | 87.8 | 8 | 12.7 | <0.001 |
| Autism subgroup |  |  |  |  |  |
| Autism with intellectual disability | 36 | 87.8 | — | — |  |
| Autism without intellectual disability | 5 | 12.2 | — | — |  |
| Consultation setting |  |  |  |  |  |
| Inpatient | 10 | 24.4 | 53 | 84.1 | <0.001 |
| Outpatient | 31 | 75.6 | 10 | 15.9 |  |
| Calendar year of ECT initiation |  |  |  |  |  |
| Median [range] | 2025<br>[2023–2026] |  | 2025<br>[2022–2026] |  | 0.756 |
| Pretreatment BFCRS total score |  |  |  |  |  |
| Mean (SD) | 16.07 (6.55) |  | 9.61 (6.75) |  | <0.001 |
| Median [IQR] | 16.0 [12.0–19.0] |  | 8.0 [5.0–13.5] |  |  |

*a Percentages were calculated among patients with evaluable pretreatment data.* *b Between-cohort p-values were calculated using Mann-Whitney U tests for age at consultation, calendar year of electroconvulsive therapy initiation, and pretreatment Bush-Francis Catatonia Rating Scale total score; chi-square tests for biologic sex and consultation setting; and Fisher's exact test for intellectual disability.* *c Consultation setting was not included in the primary adjustment set because equivalent inpatient treatment pathways were not readily available to autistic patients with intellectual disability. Consultation setting was evaluated in structural-access sensitivity analyses.* *d Pretreatment Bush-Francis Catatonia Rating Scale total scores were prorated to 23 items for assessments containing at least 20 observed items.* *Abbreviations: BFCRS, Bush-Francis Catatonia Rating Scale; ECT, electroconvulsive therapy; IQR, interquartile range; SD, standard deviation.*

Outpatient consultation occurred in 28 of 36 patients with autism and intellectual disability, 3 of 5 without intellectual disability, and 10 of 63 non-autistic patients. Clinical group was strongly associated with consultation setting (χ²=37.69, df=2, p<0.001; Cramér’s V=0.602), reflecting structurally non-equivalent pathways to ECT treatment.

### Baseline BFCRS Catatonic Phenotype

In unadjusted binary-cohort comparisons, autistic patients had higher prevalence of several activated, repetitive, and behaviorally dysregulated signs, whereas immobility/stupor and withdrawal were more prevalent in the non-autism cohort (Table 2; Figure 1A).

**Figure 1.**
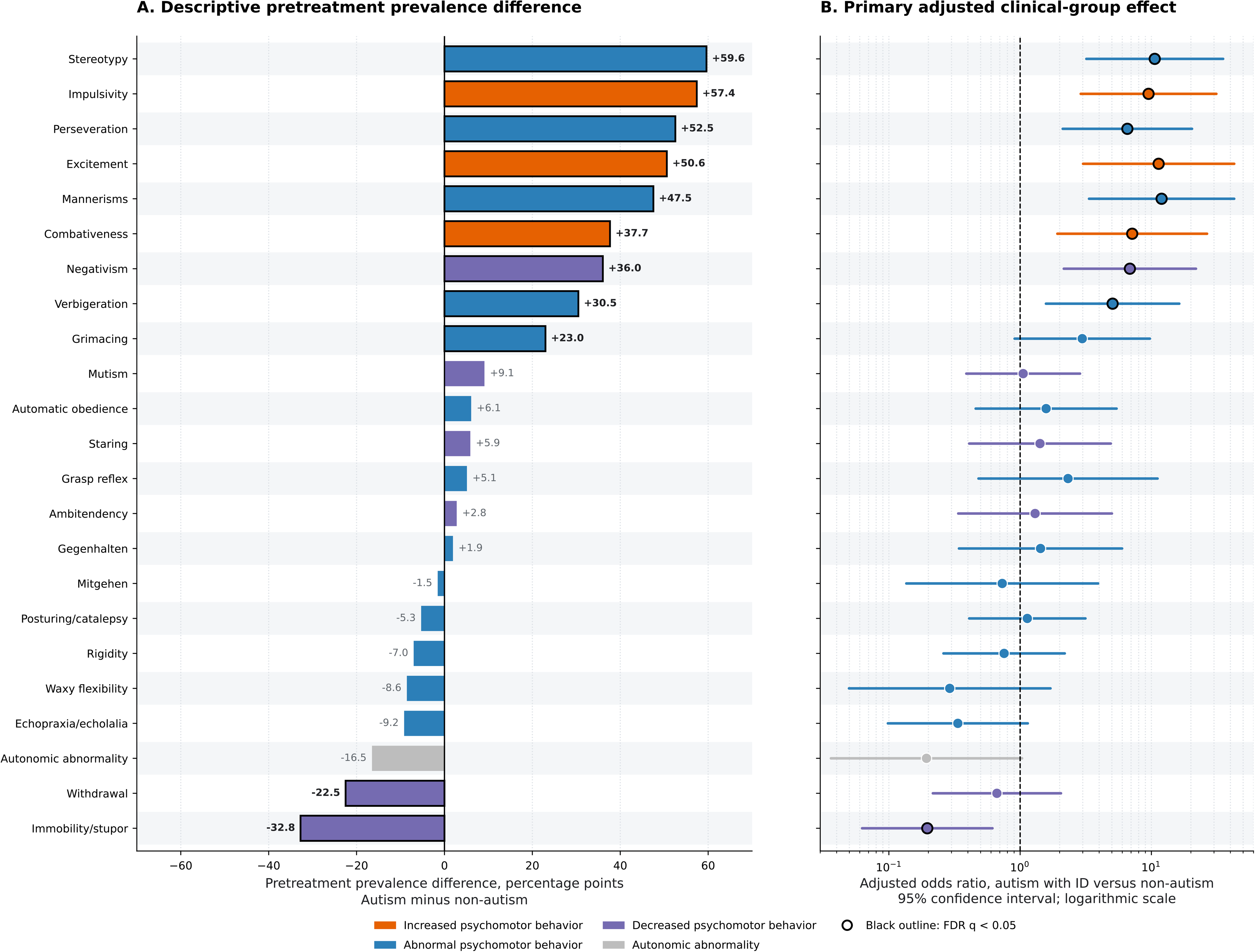
Pretreatment catatonia phenotype in autism with intellectual disability. Panel A shows unadjusted differences in the pretreatment prevalence of each of the 23 Bush-Francis Catatonia Rating Scale (BFCRS) items between autistic and non-autistic patients. Differences are expressed in percentage points and calculated as autism minus non-autism, such that positive values indicate higher prevalence in autistic patients. Panel B shows adjusted odds ratios and 95% confidence intervals comparing patients with autism and intellectual disability with the non-autism reference group. Models adjusted for age at consultation, biologic sex, and calendar year of electroconvulsive therapy initiation. The odds-ratio axis is logarithmic. Colors indicate increased, abnormal, and decreased psychomotor-behavior domains according to the checklist presented by Walther and colleagues; autonomic abnormality is shown separately because it is not a psychomotor sign. In Panel A, black outlines identify prevalence differences that remained significant after Benjamini-Hochberg FDR correction across the 23 unadjusted item comparisons. In Panel B, black outlines identify autism-with-intellectual-disability versus non-autism contrasts that remained significant after FDR correction across the 46 prespecified adjusted clinical-group contrasts. Estimates for autism without intellectual disability, consultation-setting sensitivity analyses, and flexible-age sensitivity analyses are reported in Supplemental Table 3.

**Table 2:**
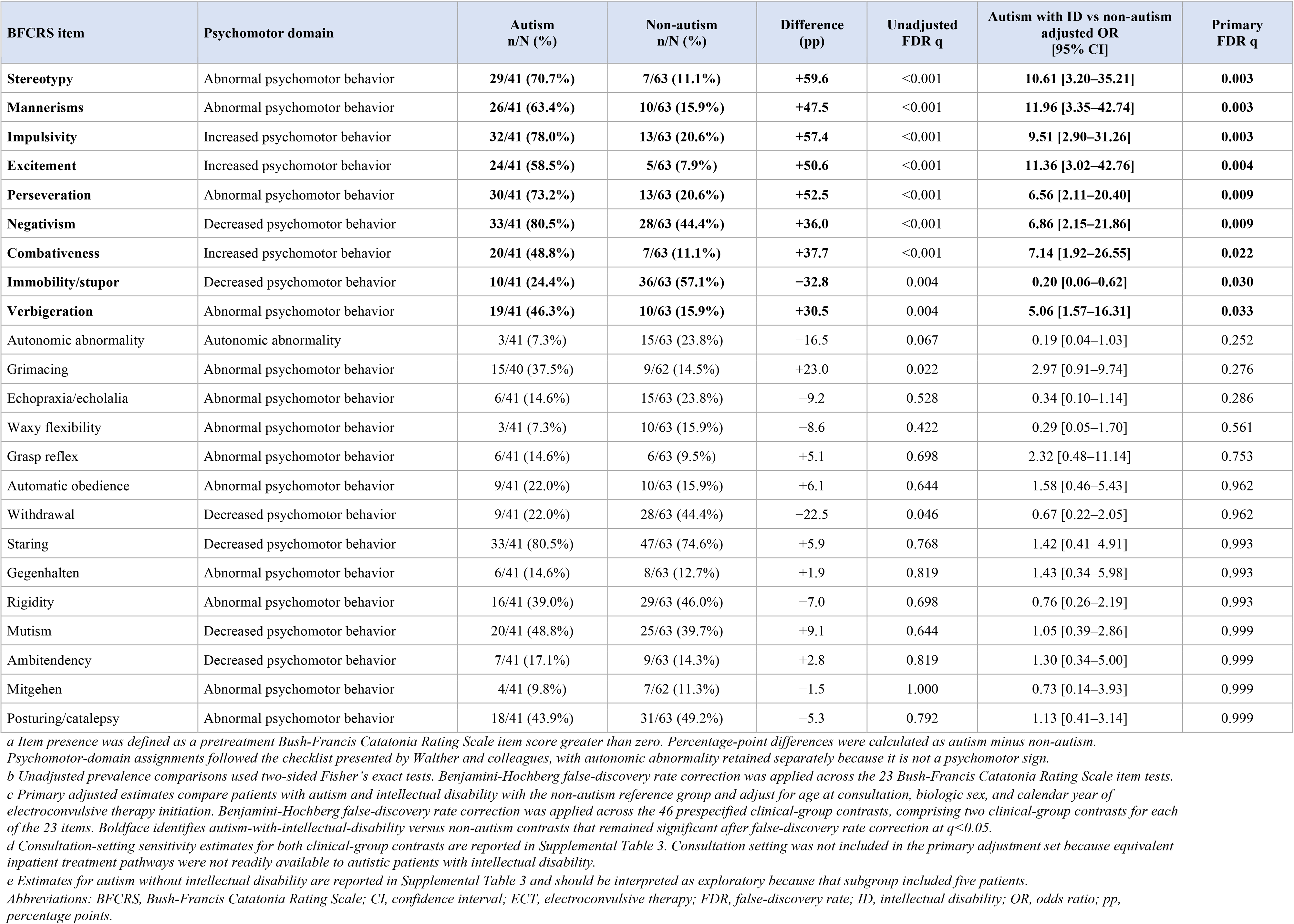
Pretreatment BFCRS Item-Level Phenotype in Autistic and Non-Autistic Patients.

In primary models using the three-level clinical group and adjusting for age, biologic sex, and calendar year of ECT initiation, nine autism-with-intellectual-disability versus non-autism BFCRS item contrasts survived FDR correction. Compared with non-autistic patients, patients with autism and intellectual disability had higher odds of stereotypy (adjusted OR=10.61, 95% CI 3.20–35.21; q=0.003), mannerisms (OR=11.96, 95% CI 3.35–42.74; q=0.003), impulsivity (OR=9.51, 95% CI 2.90–31.26; q=0.003), excitement (OR=11.36, 95% CI 3.02–42.76; q=0.004), negativism (OR=6.86, 95% CI 2.15–21.86; q=0.009), perseveration (OR=6.56, 95% CI 2.11–20.40; q=0.009), combativeness (OR=7.14, 95% CI 1.92–26.55; q=0.022), and verbigeration (OR=5.06, 95% CI 1.57–16.31; q=0.033) (Figure 1B). Immobility/stupor was less likely in autism with intellectual disability (OR=0.20, 95% CI 0.06–0.62; q=0.030).

Flexible modeling of age preserved all eight FDR-significant higher-odds associations; lower odds of immobility/stupor remained directionally consistent but did not survive FDR correction in this sensitivity model (OR=0.21, 95% CI 0.06–0.70; q=0.057; Supplemental Table 3). No autism-without-intellectual-disability contrast survived FDR correction. Estimates for this five-patient subgroup were imprecise, and several items had zero-event cells; these estimates were therefore treated as exploratory.

Additional adjustment for consultation setting attenuated the clinical-group associations, and no autism-with-intellectual-disability contrast survived correction across the setting-adjusted family. Nevertheless, effect directions were preserved for 21 of 23 items, and the principal estimates remained directionally consistent. Because consultation setting reflected structurally different care pathways, these models were interpreted as sensitivity analyses rather than definitive adjustment (Supplemental Table 3).

### BFCRS Item-Level Treatment Response

Fixed first-to-last BFCRS data were available for 96 patients, including 39 autistic and 57 non-autistic patients. Both cohorts demonstrated broadly favorable item-level change, with the largest paired effects involving increased, repetitive, and behaviorally dysregulated signs in autistic patients and hypokinetic, abnormal, and examiner-elicited signs in non-autistic patients (Table 3; Figure 2). Item-specific denominators, rank-biserial correlations, and improved, unchanged, and worsened counts are reported in Table 3. These within-patient changes do not establish that ECT alone caused the improvements because concurrent interventions were not standardized and no untreated control group was available.

**Figure 2.**
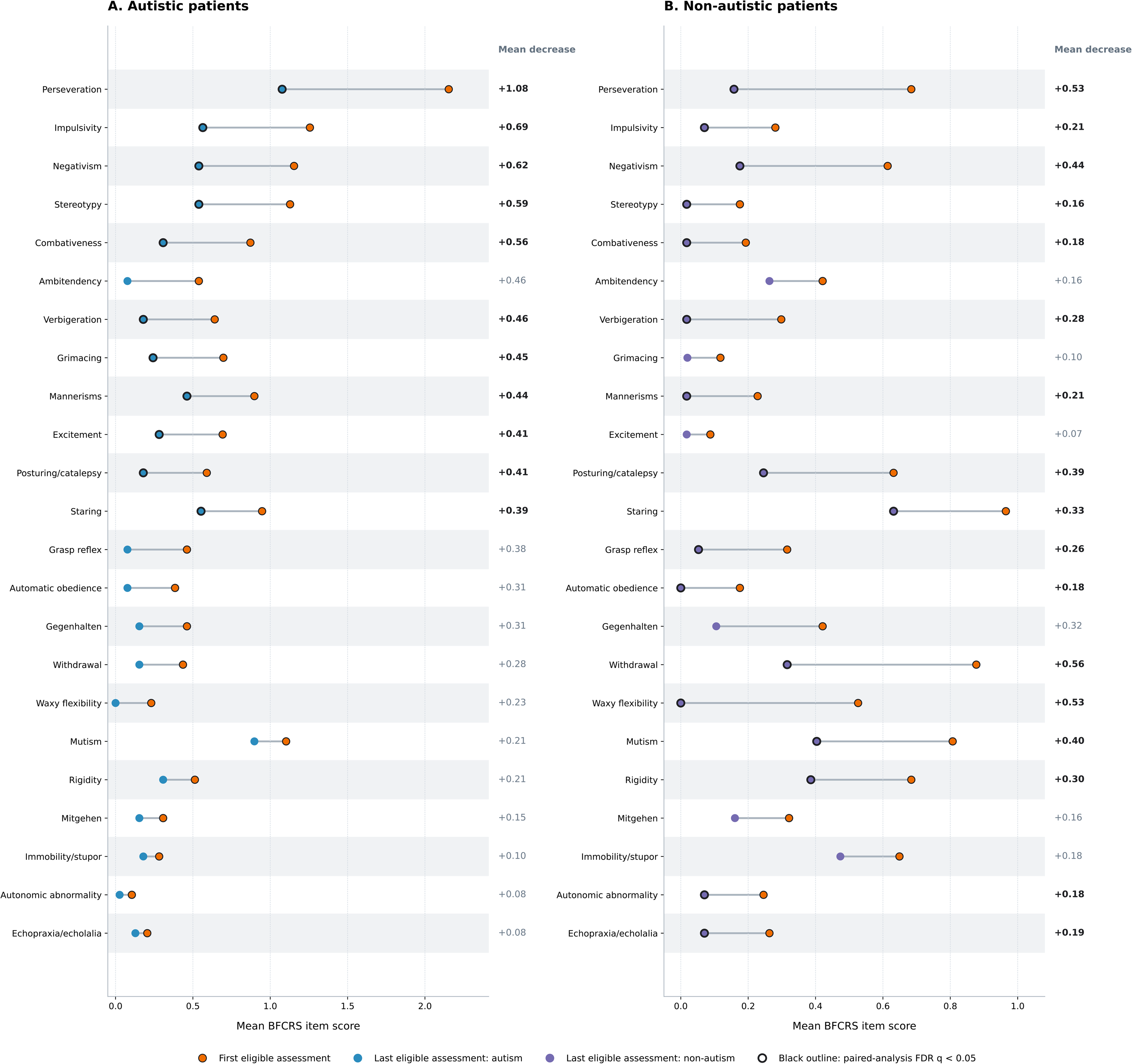
Bush-Francis Catatonia Rating Scale item-level change during electroconvulsive therapy by cohort. Items are ordered by descending mean decrease in the autism cohort using unrounded values, and the same item order is retained in the non-autism panel to facilitate direct between-panel comparison. Panel A shows mean item scores at the first and last eligible assessments among 39 autistic patients with paired BFCRS data. Panel B shows corresponding mean item scores among 57 non-autistic patients. Each panel uses its own x-axis scale. First and last eligible assessment occasions were selected at the scale level before item-level testing. All displayed first and last means and mean decreases were calculated among the item-specific patients with non-missing values at both fixed endpoints; item-specific denominators therefore varied. Mean decrease was calculated as the first score minus the last score, such that positive values indicate lower scores at the final assessment. Within each cohort, first and last item scores were compared using two-sided paired Wilcoxon signed-rank tests with Pratt handling of zero-difference ties. Black outlines identify items that remained significant after Benjamini-Hochberg FDR correction within the respective 23-item cohort-specific family at q<0.05. Lines connect group-level means and do not represent individual patient trajectories. Paired rank-biserial correlations and improved, unchanged, and worsened counts are reported in Table 3.

**Table 3:**
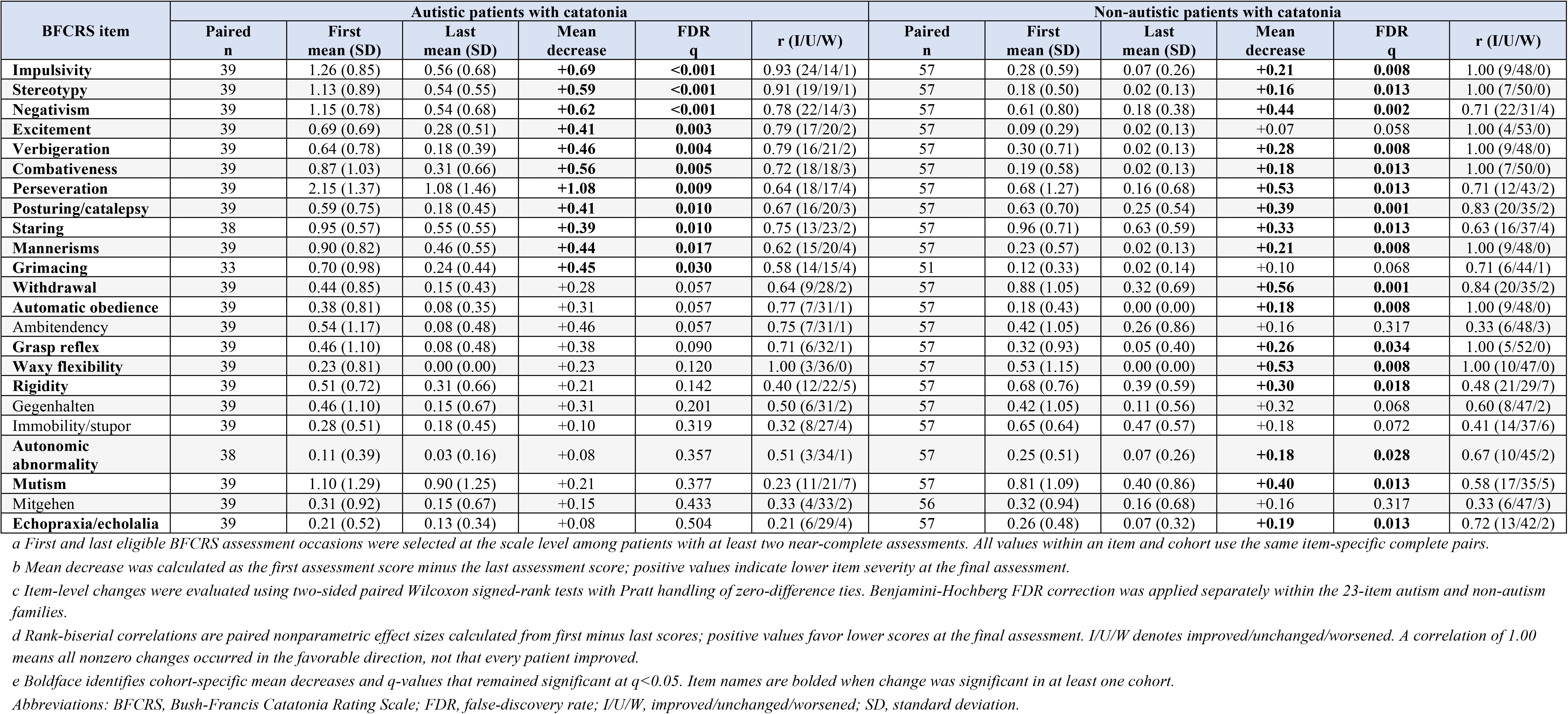
BFCRS Item-Level Change During Electroconvulsive Therapy by Cohort.

### BFCRS Secondary Longitudinal and Sensitivity Analyses

Across all 104 patients, 993 near-complete BFCRS assessments were available. Longitudinal models were restricted to the 96 patients with at least two eligible assessments, leaving 985 assessments; outcome-specific models included 983 to 985 assessments because of item-level missingness (Supplemental Table 4). Among patients with autism and intellectual disability, increased, abnormal, and decreased psychomotor-behavior scores declined over time, as did total BFCRS score, with all four principal slopes remaining significant after FDR correction.

Abnormal psychomotor-behavior scores declined (β=−0.072, 95% CI −0.098 to −0.045; q<0.001), as did decreased psychomotor-behavior scores (β=−0.060, 95% CI −0.080 to −0.040; q<0.001). Among non-autistic patients, abnormal psychomotor-behavior scores declined (β=−0.055, 95% CI −0.086 to −0.025; q<0.001), as did decreased psychomotor-behavior scores (β=−0.077, 95% CI −0.122 to −0.031; q=0.001). Increased psychomotor behavior did not decline significantly in the unrestricted non-autism model, and autonomic abnormality did not show significant unrestricted change in either group after FDR correction. Total BFCRS scores declined in autism with intellectual disability (β=−1.555, 95% CI −2.027 to −1.084; q<0.001) and non-autism (β=−1.269, 95% CI −2.004 to −0.535; q=0.001). No clinical-group-by-time interaction survived FDR correction for any psychomotor domain or total BFCRS score. Adding consultation setting produced negligible changes in the longitudinal estimates and did not alter the interaction conclusions (Supplemental Table 4).

Findings were consistent within common 30-, 60-, 90-, and 180-day follow-up windows. Total BFCRS and increased, abnormal, and decreased psychomotor-behavior scores declined in both the autism-with-intellectual-disability and non-autism groups at every window, with all corresponding principal slopes remaining significant after FDR correction. Within the 30-day window, decreased psychomotor-behavior scores declined in autism with intellectual disability (β=−0.078, 95% CI −0.149 to −0.007; p=0.031; q=0.039) and non-autism (β=−0.135, 95% CI −0.194 to −0.076; p<0.001; q<0.001). No window-specific clinical-group-by-time interaction survived FDR correction (Supplemental Table 4).

Complete-case analyses of 891 assessments from 94 patients preserved all principal coefficient directions and nominal significance classifications. Under an independence working-correlation structure, the three psychomotor-domain and total-BFCRS slopes remained negative in autism with intellectual disability, although several non-autism slopes were less robust. No principal clinical-group-by-time interaction survived FDR correction under either working-correlation specification.

### Autism-Specific Kanner Catatonia Rating Scale Analyses

KCRS analyses included 454 KCS assessments from 38 patients and 458 KCE assessments from 40 patients. Fixed endpoints were available for 35 KCS and 36 KCE patients (Supplemental Table 1; Supplemental Figure 1).

Seven KCS items decreased after FDR correction in fixed-endpoint analyses: stereotypy, staring, negativism, excitement, grimacing, impulsivity, and rigidity (Supplemental Table 1; Supplemental Figure 1). Longitudinal analyses produced a closely concordant pattern, with stereotypy, staring, negativism, excitement, impulsivity, and rigidity significant using both approaches; posturing was significant only longitudinally and grimacing only in the paired analysis (Supplemental Tables 1–2).

Aggregate Kanner scale and catatonia-related self-injury outcomes from this cohort are reported in the companion treatment-course analysis (27). The present analyses extended those findings by identifying the individual KCS and KCE features that changed during treatment.

No KCE item survived FDR correction in fixed-endpoint analyses (Supplemental Table 1; Supplemental Figure 1). Perseveration and command-verbal behavior showed the largest descriptive reductions, but both had q=0.070; other item-level estimates are reported in Supplemental Table 1. In sparsity-screened longitudinal analyses, five of eight eligible KCE items showed declining odds over time after FDR correction, although inference was limited by low event counts (Supplemental Table 2).

Overall, KCS findings provided the strongest autism-specific item-level evidence of treatment-associated improvement, whereas KCE inference was constrained by sparse events.

## Discussion

To our knowledge, this is the largest item-level comparison of BFCRS phenotype in autistic and non-autistic patients with catatonia, building on prior systematic reviews of smaller studies (18,20). Nine autism-with-intellectual-disability versus non-autism contrasts survived FDR correction: eight activated, repetitive, or behaviorally dysregulated signs had higher adjusted odds, whereas immobility/stupor had lower adjusted odds. These signs overlap with behaviors that may be attributed to autism itself, particularly in individuals with intellectual disability or profound autism (8,18,20,41,42). Their emergence or substantial worsening, together with treatment-associated improvement, supports their interpretation as potential manifestations of catatonia in appropriately selected patients rather than as inherent features of autism (17).

This phenotype did not correspond to a single Walther psychomotor domain (39). It included all three increased-domain signs, repetitive and verbal signs from the abnormal domain, and negativism from the decreased domain. In contrast, the classically hypokinetic sign immobility/stupor was less prevalent in autism with intellectual disability. The phenotype should therefore be interpreted as a descriptive clinical grouping spanning established psychomotor domains, not as an independently validated catatonia subtype.

In practice, catatonia may create the appearance of worsening autism, making an autistic individual seem more impaired or developmentally regressed than their pre-catatonia baseline and obscuring recognition of a superimposed, potentially treatable syndrome. This distinction is especially important during later regression, which differs from the early-childhood regression traditionally associated with autism (17,23–25). Early-childhood autistic regression most commonly involves loss or decline in language or social-communication skills during the second year of life, whereas later regression occurs after a period of relatively stable functioning (23,24). In later regression, the diagnostic signal is not the presence of a behavior alone, but a meaningful departure from the individual’s baseline in its frequency, intensity, functional consequences, or effects on medical stability. An autistic individual may have longstanding stereotypy, limited spoken communication, or dependence on others for daily activities; however, the emergence or substantial worsening of these features may reflect catatonia rather than progression of autism. New or progressive loss of communication, self-care, mobility, continence, food intake, behavioral regulation, or previously acquired skills should therefore prompt evaluation for catatonia and other potentially treatable causes.

The July 2026 Interagency Autism Coordinating Committee (IACC) Strategic Plan working draft identifies neurodevelopmental regression as an area requiring coordinated research and proposes a National Neurodevelopmental Regression Initiative (43). The present findings align with this emerging priority by illustrating the need for longitudinal phenotyping and timely evaluation of potentially treatable causes of regression, including catatonia; however, the draft is not an adopted federal recommendation.

Treatment-associated improvement was evident at item, psychomotor-domain, and total-score levels. Increased, abnormal, and decreased psychomotor behavior declined over time in patients with autism and intellectual disability, while abnormal and decreased psychomotor behavior declined over time in non-autistic patients. Within the common follow-up windows, increased, abnormal, and decreased psychomotor-behavior scores declined in both groups at 30, 60, 90, and 180 days, with these principal group-specific findings remaining significant after FDR correction. No clinical-group-by-time interaction survived FDR correction. However, the absence of a significant interaction does not establish equivalent treatment effects, and the observational design does not establish that ECT alone caused the changes. These findings were robust to complete-case outcome construction, while some unrestricted non-autism slopes were sensitive to the working-correlation specification.

The companion analysis modeled change per ECT session and found more gradual BFCRS decline in autism with intellectual disability, whereas the present models evaluated change over log-transformed elapsed time and identified no FDR-significant differences in psychomotor-domain trajectories. These analyses estimate different quantities, namely symptom change per delivered treatment versus change across observed clinical time. Differences in treatment frequency, maintenance duration, assessment timing, and outcome construction may therefore account for the differing interaction results (27).

In a companion analysis of the same cohort (27), we found that autistic patients received more ECT sessions over longer treatment durations (median 35 vs 14 sessions; 342 vs 97 days). The present analysis complements those treatment-burden findings by identifying the catatonic features present at consultation and the features that changed during treatment. It does not establish that activated or repetitive signs themselves require longer treatment, because treatment duration may reflect baseline severity, intellectual disability, relapse risk, maintenance needs, treatment access, and other clinical or structural factors. Nevertheless, the item-level and longitudinal findings demonstrate that behaviors easily misattributed to autism were not invariably fixed and showed treatment-associated improvement. Several features associated with autism and intellectual disability in this study, particularly combativeness, impulsivity, stereotypy, and mannerisms, overlap with presentations historically described as treatment-refractory aggression or self-injury (36,44–46). When these behaviors are new, markedly worsened, or accompanied by loss of communication, self-care, mobility, nutrition, continence, or broader functional regression, catatonia should be considered alongside psychiatric, neurologic, genetic, medical, environmental, and behavioral explanations. Recognition of catatonia does not negate the underlying autism diagnosis. It identifies a potentially reversible syndrome superimposed on a chronic neurodevelopmental condition.

These observations raise a broader question about the neurobiological relationship between catatonia and autism. The motor and behavioral features that define catatonia, including stereotypy, perseveration, negativism, impulsivity, and echolalia, are also core or associated features of autism spectrum disorder, particularly in individuals with co-occurring intellectual disability or profound autism. Whether this phenotypic overlap reflects shared neural circuitry, such as dysfunction in cortico-striatal-thalamic loops implicated in both conditions (39), or whether catatonia represents a distinct pathophysiological state that preferentially recruits motor circuits already vulnerable in autism remains an open question. Recently, this question was posed by Mahgoub and colleagues in a review of the literature. In their review, they found weak support for separating catatonia from overlapping restricted interests and repetitive behaviors (47), suggesting that some core features of autism may align more with catatonia than features intrinsic to autism. Although the present data cannot resolve this question, the observation that catatonic symptoms phenotypically overlapping with autism improved during ECT while the underlying autism diagnosis persisted is consistent with catatonia acting as a potentially reversible regressive exacerbation superimposed on a chronic neurodevelopmental substrate.

One possible explanation for the differing catatonia phenotypes is that psychomotor disturbances are expressed differently across neurobiological substrates. Parvalbumin-interneuron abnormalities, altered cortical modulation, and excitation-inhibition imbalance have been proposed in autism, particularly among individuals with cognitive impairment (48–54), while GABAergic and motor-network dysfunction have been implicated in catatonia (55). Language and verbal behavior may represent one dimension of this heterogeneity. Wing and Shah found that autistic individuals with later severe catatonic deterioration were more likely to have had impaired language before behavioral change (11). In a pilot TMS biomarker study, greater caregiver-rated inappropriate speech, reflecting excessive, repetitive, or atypical verbal output rather than language ability, was associated with progressively greater post-cTBS M1 facilitation in autistic participants (56). Catatonia severity did not significantly moderate the post-cTBS response, although interpretation was limited by low symptom burden and a small, procedurally selected sample. These distinct findings do not establish a shared mechanism but suggest that reduced language capacity and increased atypical verbal output may identify heterogeneity in motor, speech, and behavioral-network regulation. Whether such differences influence activated, repetitive, or hypokinetic catatonia presentations requires direct neurophysiological study in patients with clinically significant catatonia.

This study has several limitations. First, it was conducted at a single tertiary academic center with a specialized ECT program for neurodevelopmental disorders. The referred autism cohort may overrepresent individuals with intellectual disability, severe catatonia, or treatment-refractory illness. Only five autistic patients did not have intellectual disability, making estimates for that subgroup exploratory and imprecise.

Second, clinical group and consultation setting were strongly associated because equivalent inpatient ECT pathways were not readily available to autistic patients with intellectual disability. Consultation setting could represent structural access, referral selection, and clinical presentation simultaneously. Primary models therefore excluded setting from the conventional adjustment set, while setting-adjusted estimates were reported as sensitivity analyses. This strategy improves transparency but cannot fully separate phenotype from referral structure.

Third, follow-up duration and treatment intensity differed substantially between cohorts. Longitudinal models incorporated elapsed time and repeated assessments, and conclusions were consistent across 30-, 60-, 90-, and 180-day common follow-up windows. These approaches nevertheless cannot eliminate selection associated with treatment continuation or maintenance ECT.

Fourth, clinical ratings were collected during routine care by raters who were not blinded to diagnosis or treatment course, and inter-rater reliability was not assessed. Concurrent medications, behavioral interventions, and other clinical changes were not standardized. The analyses therefore characterize treatment-associated within-patient change and do not establish that ECT alone caused the observed improvements.

Fifth, eight patients with pretreatment BFCRS data did not have a second eligible assessment and were excluded from fixed-endpoint analyses. The BFCRS has not been formally validated specifically in autistic or pediatric catatonia, and higher scores in autistic patients may partly reflect overlap between catatonic deterioration and longstanding neurodevelopmental features. Determining change from pre-catatonia functioning remains essential. Although later regression provides an important clinical framework for interpreting these findings, regression was not systematically measured across the cohort. This study could not consistently quantify its timing, tempo, affected functional domains, precipitating factors, or relationship to treatment initiation. Therefore, regression cannot be treated as a formal exposure or outcome in this analysis, and the findings should not be interpreted as estimating the prevalence of catatonia among autistic individuals with regression.

Finally, KCS and KCE assessments were available only in autistic patients and could not support between-cohort comparisons. Kanner follow-up intervals were heterogeneous, although longitudinal models incorporated elapsed time and repeated observations. KCE signs were often uncommon; no fixed-endpoint KCE item survived FDR correction, and several longitudinal models were excluded because of sparse events or insufficient within-patient variation.

In conclusion, catatonia in patients with autism and intellectual disability was characterized by an activated, repetitive, and behaviorally dysregulated phenotype, whereas immobility/stupor, a classically hypokinetic sign, was more prominent in non-autistic catatonia in the primary analysis. Many of the features associated with autism and intellectual disability overlap with behaviors commonly used to judge autism severity and functional status. Consequently, superimposed catatonia may make an autistic individual appear more impaired or regressed than their baseline, obscuring recognition of a potentially reversible syndrome. Catatonia should be considered when autistic individuals experience later behavioral, motor, or functional regression, particularly when stereotypy, perseveration, impulsivity, combativeness, excitement, self-injury, communication, mobility, self-care, or food intake changes substantially from prior functioning. Screening instruments such as the Catatonia Quick Screen may support case identification but do not replace a comprehensive evaluation of change from baseline and potentially treatable causes of regression (57,58). Early recognition and treatment of catatonia in this population may reduce morbidity and improve long-term outcomes for autistic individuals and their families.

## Supporting information

STROBE checklist

## Author Contributions

Joshua Ryan Smith (JRS) conceptualized the study and led the methodology, formal analysis, investigation, and data curation; JRS also wrote the original draft and provided overall supervision. Maria Bonnee (MB) contributed to data curation and investigation. Seri Lim (SL) contributed to data curation and investigation. D. Catherine Fuchs (CF) provided supervision and contributed to manuscript review and editing. Sarah Marler (SM) provided supervision and contributed to manuscript review and editing. Isaac Baldwin (IB) contributed to conceptualization, investigation, data collection, and manuscript review and editing. Rafael Tamargo (RT), Christopher Maley (CM), Ashley VanHaverbeck (AV), Courtney Hamilton (CH), and Timothy Adegoke (TA) contributed to data collection and manuscript review and editing. Haozheng Xu (HX) contributed to data collection, formal analysis, and manuscript review and editing. Zachary J. Williams (ZJW) contributed to conceptualization, investigation, and manuscript review and editing. Jo Ellen Wilson (JEW) contributed to conceptualization, supervision, and manuscript review and editing. James Luccarelli (JL) contributed to conceptualization, investigation, supervision, and manuscript review and editing. All authors reviewed and approved the final manuscript.

## Data Availability

De-identified data for this study are available upon reasonable request.

## Ethical approval for human patients

Informed consent for electroconvulsive therapy was obtained for all patients in accordance with Tennessee state law. Informed consent for participation in this observational study was waived by the Vanderbilt University Medical Center Institutional Review Board (Number: 242126) because data were collected as part of routine clinical care.

## Abbreviations

ECT: Electroconvulsive therapy
BFCRS: Bush-Francis Catatonia Rating Scale
FDR: false-discovery rate
KCRS: Kanner Catatonia Rating Scale
KCS: Kanner Catatonia Severity scale
KCE: Kanner Catatonia Examination
STROBE: Strengthening the Reporting of Observational Studies in Epidemiology
VUMC: Vanderbilt University Medical Center
DSM-5: Diagnostic and Statistical Manual of Mental Disorders, Fifth Edition
GEE: generalized estimating equations
IACC: Interagency Autism Coordinating Committee

---

**Supplemental Figure 1. Autism-specific Kanner item-level change during treatment.** Panel A shows mean Kanner Catatonia Severity (KCS) item scores at the first and last eligible assessments among 35 autistic patients with paired KCS data. Positive mean decreases represent lower item scores at the last assessment. Panel B shows the percentage of patients with each Kanner Catatonia Examination (KCE) sign present at the first and last eligible assessments among 36 autistic patients with paired KCE data. Negative prevalence changes represent lower prevalence at the last assessment. Black outlines identify items that remained significant after Benjamini-Hochberg false-discovery rate correction within the respective 18-item KCS and 12-item KCE test families. KCS changes were evaluated using two-sided paired Wilcoxon signed-rank tests with Pratt handling of zero-difference ties. KCE changes were evaluated using two-sided exact McNemar tests. First and last assessments were selected at the scale level before item-level testing; item-specific denominators could vary if an item was missing at either fixed endpoint. Lines connect group-level first and last values and do not represent individual patient trajectories.

