## Supplementary figures and images for "Catatonic Features That May Be Mistaken for Worsening Autism Show Treatment-Associated Improvement: Implications for Later Regression"

### Supplemental Figure 1

#### Supplemental Figure 1: Autism-Specific Kanner Item-Level Change

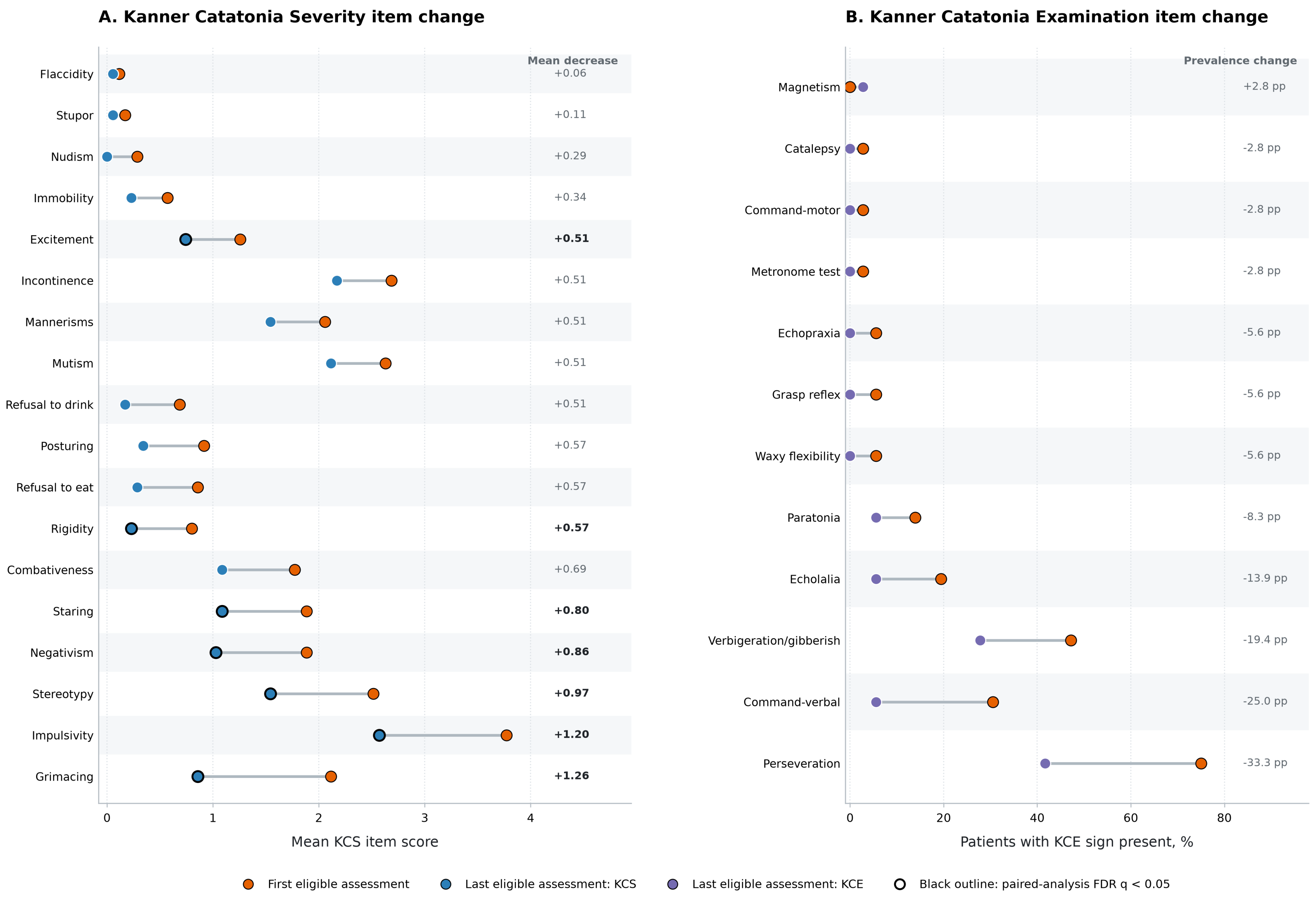
