## Supplemental Table 1 for "Catatonic Features That May Be Mistaken for Worsening Autism Show Treatment-Associated Improvement: Implications for Later Regression"

**Supplemental Table 1: Autism-Specific Kanner Item-Level Change at Fixed Endpoints**

| **Kanner Catatonia Severity (KCS)** | | | | | | | | | |
| --- | --- | --- | --- | --- | --- | --- | --- | --- | --- |
| **KCS item** | **Paired n** | **First mean (SD)** | **Last mean (SD)** | **Mean decrease** | **Improved** | **Unchanged** | **Worsened** | **Rank-biserial correlation** | **FDR q** |
| **Stereotypy** | 35 | 2.51 (1.70) | 1.54 (1.46) | **+0.97** | 15 | 18 | 2 | 0.80 | **0.023** |
| **Staring** | 35 | 1.89 (0.96) | 1.09 (1.01) | **+0.80** | 14 | 19 | 2 | 0.78 | **0.023** |
| **Negativism** | 35 | 1.89 (1.45) | 1.03 (1.32) | **+0.86** | 16 | 16 | 3 | 0.65 | **0.023** |
| **Grimacing** | 35 | 2.11 (2.74) | 0.86 (1.83) | **+1.26** | 11 | 22 | 2 | 0.85 | **0.028** |
| **Impulsivity** | 35 | 3.77 (2.41) | 2.57 (2.30) | **+1.20** | 18 | 10 | 7 | 0.61 | **0.028** |
| **Excitement** | 35 | 1.26 (1.09) | 0.74 (1.09) | **+0.51** | 10 | 24 | 1 | 0.82 | **0.028** |
| **Rigidity** | 35 | 0.80 (1.30) | 0.23 (0.81) | **+0.57** | 10 | 23 | 2 | 0.72 | **0.049** |
| Posturing | 35 | 0.91 (1.70) | 0.34 (0.76) | +0.57 | 8 | 25 | 2 | 0.67 | 0.095 |
| Refusal to eat | 35 | 0.86 (1.83) | 0.29 (1.10) | +0.57 | 6 | 28 | 1 | 0.89 | 0.095 |
| Refusal to drink | 35 | 0.69 (1.75) | 0.17 (1.01) | +0.51 | 4 | 31 | 0 | 1.00 | 0.095 |
| Combativeness | 35 | 1.77 (2.16) | 1.09 (1.48) | +0.69 | 13 | 16 | 6 | 0.46 | 0.134 |
| Mannerisms | 35 | 2.06 (2.09) | 1.54 (1.54) | +0.51 | 9 | 22 | 4 | 0.52 | 0.202 |
| Nudism | 35 | 0.29 (1.20) | 0.00 (0.00) | +0.29 | 2 | 33 | 0 | 1.00 | 0.218 |
| Mutism | 35 | 2.63 (3.21) | 2.11 (3.25) | +0.51 | 10 | 20 | 5 | 0.34 | 0.254 |
| Immobility | 35 | 0.57 (1.42) | 0.23 (0.65) | +0.34 | 5 | 28 | 2 | 0.57 | 0.286 |
| Incontinence | 35 | 2.69 (3.10) | 2.17 (3.12) | +0.51 | 11 | 18 | 6 | 0.22 | 0.310 |
| Stupor | 35 | 0.17 (0.57) | 0.06 (0.34) | +0.11 | 3 | 31 | 1 | 0.50 | 0.317 |
| Flaccidity | 35 | 0.11 (0.47) | 0.06 (0.34) | +0.06 | 1 | 34 | 0 | 1.00 | 0.317 |

| **Kanner Catatonia Examination (KCE)** | | | | | | | | |
| --- | --- | --- | --- | --- | --- | --- | --- | --- |
| **KCE item** | **Paired n** | **First present n (%)** | **Last present n (%)** | **Prevalence change (pp)** | **Resolved** | **Emerged** | **Exact p** | **FDR q** |
| Perseveration | 36 | 27 (75.0%) | 15 (41.7%) | −33.3 | 15 | 3 | 0.008 | 0.070 |
| Command-verbal | 36 | 11 (30.6%) | 2 (5.6%) | −25.0 | 10 | 1 | 0.012 | 0.070 |
| Verbigeration/gibberish | 36 | 17 (47.2%) | 10 (27.8%) | −19.4 | 8 | 1 | 0.039 | 0.156 |
| Echolalia | 36 | 7 (19.4%) | 2 (5.6%) | −13.9 | 5 | 0 | 0.062 | 0.188 |
| Paratonia | 36 | 5 (13.9%) | 2 (5.6%) | −8.3 | 3 | 0 | 0.250 | 0.600 |
| Waxy flexibility | 36 | 2 (5.6%) | 0 (0.0%) | −5.6 | 2 | 0 | 0.500 | 0.750 |
| Echopraxia | 36 | 2 (5.6%) | 0 (0.0%) | −5.6 | 2 | 0 | 0.500 | 0.750 |
| Grasp reflex | 36 | 2 (5.6%) | 0 (0.0%) | −5.6 | 2 | 0 | 0.500 | 0.750 |
| Catalepsy | 36 | 1 (2.8%) | 0 (0.0%) | −2.8 | 1 | 0 | 1.000 | 1.000 |
| Command-motor | 36 | 1 (2.8%) | 0 (0.0%) | −2.8 | 1 | 0 | 1.000 | 1.000 |
| Metronome test | 36 | 1 (2.8%) | 0 (0.0%) | −2.8 | 1 | 0 | 1.000 | 1.000 |
| Magnetism | 36 | 0 (0.0%) | 1 (2.8%) | +2.8 | 0 | 1 | 1.000 | 1.000 |

*a Fixed first and last eligible assessment occasions were selected at the scale level before item-level testing. Item-specific paired denominators could vary if an item was missing at either fixed endpoint.
b KCS mean decrease was calculated as the first assessment score minus the last assessment score; positive values indicate lower item severity at the last assessment. KCS changes were evaluated using two-sided paired Wilcoxon signed-rank tests with Pratt handling of zero-difference ties.
c KCE items were binary. Prevalence change was calculated as last-assessment prevalence minus first-assessment prevalence; negative values indicate lower prevalence at the last assessment. KCE changes were evaluated using two-sided exact McNemar tests.
d Benjamini-Hochberg FDR correction was applied separately within the 18-item KCS and 12-item KCE paired-test families. Boldface identifies KCS results that remained significant at q<0.05.
e No individual KCE item remained significant after FDR correction.
Abbreviations: FDR, false-discovery rate; KCE, Kanner Catatonia Examination; KCS, Kanner Catatonia Severity; pp, percentage points; SD, standard deviation.*
