## Supplemental Table 2 for "Catatonic Features That May Be Mistaken for Worsening Autism Show Treatment-Associated Improvement: Implications for Later Regression"

**Supplemental Table 2: Autism-Specific Longitudinal Kanner Item Models**

| **KCS item** | **Assessments** | **Patients** | **Time β** | **SE** | **95% CI** | **Raw p** | **FDR q** |
| --- | --- | --- | --- | --- | --- | --- | --- |
| **Negativism** | 454 | 38 | **-0.221** | 0.045 | [-0.310 to -0.133] | <0.001 | **<0.001** |
| **Impulsivity** | 454 | 38 | **-0.297** | 0.085 | [-0.464 to -0.130] | <0.001 | **0.004** |
| **Staring** | 454 | 38 | **-0.120** | 0.040 | [-0.199 to -0.041] | 0.003 | **0.014** |
| **Stereotypy** | 454 | 38 | **-0.155** | 0.052 | [-0.257 to -0.053] | 0.003 | **0.014** |
| **Excitement** | 454 | 38 | **-0.109** | 0.039 | [-0.185 to -0.033] | 0.005 | **0.016** |
| **Rigidity** | 454 | 38 | **-0.128** | 0.046 | [-0.218 to -0.038] | 0.005 | **0.016** |
| **Posturing** | 454 | 38 | **-0.155** | 0.065 | [-0.283 to -0.027] | 0.018 | **0.046** |
| Grimacing | 454 | 38 | -0.229 | 0.103 | [-0.430 to -0.028] | 0.026 | 0.058 |
| Combativeness | 454 | 38 | -0.138 | 0.072 | [-0.279 to 0.003] | 0.056 | 0.111 |
| Refusal to eat | 454 | 38 | -0.101 | 0.054 | [-0.207 to 0.005] | 0.063 | 0.113 |
| Refusal to drink | 454 | 38 | -0.087 | 0.050 | [-0.184 to 0.010] | 0.079 | 0.121 |
| Immobility | 454 | 38 | -0.088 | 0.051 | [-0.188 to 0.011] | 0.081 | 0.121 |
| Stupor | 454 | 38 | -0.035 | 0.024 | [-0.082 to 0.012] | 0.147 | 0.204 |
| Incontinence | 454 | 38 | -0.152 | 0.110 | [-0.368 to 0.063] | 0.166 | 0.214 |
| Nudism | 454 | 38 | -0.042 | 0.033 | [-0.107 to 0.022] | 0.197 | 0.236 |
| Flaccidity | 454 | 38 | 0.018 | 0.016 | [-0.013 to 0.049] | 0.263 | 0.286 |
| Mannerisms | 454 | 38 | -0.074 | 0.067 | [-0.205 to 0.057] | 0.270 | 0.286 |
| Mutism | 454 | 38 | -0.062 | 0.102 | [-0.261 to 0.137] | 0.543 | 0.543 |

| **KCE item** | **Assessments** | **Patients** | **Positive observations** | **Variable patients, n** | **Eligible** | **Time OR** | **95% CI** | **Raw p** | **FDR q** |
| --- | --- | --- | --- | --- | --- | --- | --- | --- | --- |
| **Command-verbal** | 458 | 40 | 59 | 18 | Yes | **0.695** | [0.596–0.811] | <0.001 | **<0.001** |
| **Paratonia** | 458 | 40 | 27 | 11 | Yes | **0.745** | [0.614–0.905] | 0.003 | **0.012** |
| **Verbigeration/gibberish** | 458 | 40 | 141 | 18 | Yes | **0.770** | [0.635–0.933] | 0.008 | **0.020** |
| **Catalepsy** | 458 | 40 | 10 | 6 | Yes | **0.776** | [0.631–0.953] | 0.016 | **0.031** |
| **Magnetism** | 458 | 40 | 10 | 7 | Yes | **0.761** | [0.599–0.965] | 0.024 | **0.039** |
| Echolalia | 458 | 40 | 35 | 10 | Yes | 0.809 | [0.656–0.998] | 0.047 | 0.063 |
| Perseveration | 458 | 40 | 301 | 22 | Yes | 0.867 | [0.734–1.025] | 0.095 | 0.108 |
| Echopraxia | 458 | 40 | 10 | 6 | Yes | 0.808 | [0.566–1.154] | 0.242 | 0.242 |
| Waxy flexibility | 458 | 40 | 11 | 4 | No | — | — | — | — |
| Grasp reflex | 458 | 40 | 4 | 2 | No | — | — | — | — |
| Command-motor | 458 | 40 | 3 | 2 | No | — | — | — | — |
| Metronome test | 458 | 40 | 1 | 1 | No | — | — | — | — |

a *Kanner Catatonia Severity Gaussian generalized estimating equation models used all eligible assessments, an identity link, patient-level clustering, an exchangeable working-correlation structure, robust standard errors, and adjustment for age at consultation, biologic sex, and calendar year of electroconvulsive therapy initiation. Time was represented as ln(days+1) from the first eligible Kanner assessment. Negative β coefficients indicate declining item severity over time.*

b *Post hoc Kanner Catatonia Examination longitudinal models used binomial generalized estimating equations with a logit link. Models were eligible only when an item had at least 10 positive observations, at least 10 negative observations, and at least five patients with within-patient variation. “Variable patients” indicates the number of patients whose item status changed across assessments.*

c *Kanner Catatonia Examination time odds ratios below 1 indicate declining odds of the examination sign with increasing ln(days+1).*

d *Benjamini-Hochberg false-discovery rate correction was applied separately across the 18 Kanner Catatonia Severity models and the eight eligible, successfully fitted Kanner Catatonia Examination models. Boldface identifies estimates that remained significant at q<0.05.*

e *Catalepsy and magnetism each had only 10 positive observations and should be interpreted cautiously despite meeting the prespecified sparsity screen.*

***Abbreviations:*** *CI, confidence interval; ECT, electroconvulsive therapy; FDR, false-discovery rate; GEE, generalized estimating equation; KCE, Kanner Catatonia Examination; KCS, Kanner Catatonia Severity; OR, odds ratio; SE, standard error.*
