## Supplemental Table 3 for "Catatonic Features That May Be Mistaken for Worsening Autism Show Treatment-Associated Improvement: Implications for Later Regression"

**Supplemental Table 3. Full Adjusted BFCRS Contrasts and Sensitivity Analyses**

| **BFCRS item-level adjusted contrasts** | | | | | | | | |
| --- | --- | --- | --- | --- | --- | --- | --- | --- |
| **BFCRS item** | **Contrast** | **Primary-adjusted OR [95% CI]** | **Primary p** | **Primary FDR q** | **Setting-adjusted OR [95% CI]** | **Setting p** | **Setting FDR q** | **Direction preserved** |
| **Stereotypy** | Autism with ID vs non-autism | **10.61 [3.20–35.21]** | <0.001 | **0.003** | 5.09 [1.29–20.05] | 0.020 | 0.170 | Yes |
| **Mannerisms** | Autism with ID vs non-autism | **11.96 [3.35–42.74]** | <0.001 | **0.003** | 7.90 [2.04–30.59] | 0.003 | 0.126 | Yes |
| **Impulsivity** | Autism with ID vs non-autism | **9.51 [2.90–31.26]** | <0.001 | **0.003** | 5.53 [1.53–20.00] | 0.009 | 0.141 | Yes |
| **Excitement** | Autism with ID vs non-autism | **11.36 [3.02–42.76]** | <0.001 | **0.004** | 7.40 [1.80–30.37] | 0.005 | 0.126 | Yes |
| **Negativism** | Autism with ID vs non-autism | **6.86 [2.15–21.86]** | 0.001 | **0.009** | 3.67 [1.03–13.03] | 0.044 | 0.255 | Yes |
| **Perseveration** | Autism with ID vs non-autism | **6.56 [2.11–20.40]** | 0.001 | **0.009** | 4.15 [1.20–14.36] | 0.025 | 0.170 | Yes |
| **Combativeness** | Autism with ID vs non-autism | **7.14 [1.92–26.55]** | 0.003 | **0.022** | 5.40 [1.30–22.50] | 0.021 | 0.170 | Yes |
| **Immobility/stupor** | Autism with ID vs non-autism | **0.20 [0.06–0.62]** | 0.005 | **0.030** | 0.24 [0.07–0.84] | 0.026 | 0.170 | Yes |
| **Verbigeration** | Autism with ID vs non-autism | **5.06 [1.57–16.31]** | 0.007 | **0.033** | 2.88 [0.79–10.42] | 0.108 | 0.551 | Yes |
| Autonomic abnormality | Autism with ID vs non-autism | 0.19 [0.04–1.03] | 0.055 | 0.252 | 0.40 [0.06–2.47] | 0.322 | 0.865 | Yes |
| Grimacing | Autism with ID vs non-autism | 2.97 [0.91–9.74] | 0.072 | 0.276 | 2.09 [0.55–7.88] | 0.279 | 0.854 | Yes |
| Echopraxia/echolalia | Autism with ID vs non-autism | 0.34 [0.10–1.14] | 0.081 | 0.286 | 0.38 [0.10–1.54] | 0.176 | 0.677 | Yes |
| Waxy flexibility | Autism with ID vs non-autism | 0.29 [0.05–1.70] | 0.171 | 0.561 | 0.29 [0.04–2.01] | 0.211 | 0.746 | Yes |
| Grasp reflex | Autism with ID vs non-autism | 2.32 [0.48–11.14] | 0.294 | 0.753 | 2.69 [0.45–16.14] | 0.278 | 0.854 | Yes |
| Automatic obedience | Autism with ID vs non-autism | 1.58 [0.46–5.43] | 0.470 | 0.962 | 3.09 [0.70–13.64] | 0.137 | 0.574 | Yes |
| Withdrawal | Autism with ID vs non-autism | 0.67 [0.22–2.05] | 0.477 | 0.962 | 0.78 [0.22–2.75] | 0.695 | 0.999 | Yes |
| Staring | Autism with ID vs non-autism | 1.42 [0.41–4.91] | 0.582 | 0.993 | 1.69 [0.42–6.87] | 0.463 | 0.999 | Yes |
| Rigidity | Autism with ID vs non-autism | 0.76 [0.26–2.19] | 0.605 | 0.993 | 0.75 [0.23–2.46] | 0.635 | 0.999 | Yes |
| Gegenhalten | Autism with ID vs non-autism | 1.43 [0.34–5.98] | 0.624 | 0.993 | 1.82 [0.34–9.63] | 0.480 | 0.999 | Yes |
| Ambitendency | Autism with ID vs non-autism | 1.30 [0.34–5.00] | 0.703 | 0.999 | 1.35 [0.30–6.10] | 0.696 | 0.999 | Yes |
| Mitgehen | Autism with ID vs non-autism | 0.73 [0.14–3.93] | 0.714 | 0.999 | 0.99 [0.15–6.67] | 0.994 | 0.999 | Yes |
| Posturing/catalepsy | Autism with ID vs non-autism | 1.13 [0.41–3.14] | 0.809 | 0.999 | 0.88 [0.27–2.81] | 0.823 | 0.999 | No |
| Mutism | Autism with ID vs non-autism | 1.05 [0.39–2.86] | 0.917 | 0.999 | 0.85 [0.27–2.66] | 0.785 | 0.999 | No |
| Mannerisms | Autism without ID vs non-autism | 6.85 [0.86–54.49] | 0.069 | 0.276 | 5.27 [0.62–44.75] | 0.128 | 0.574 | Yes |
| Impulsivity | Autism without ID vs non-autism | 3.12 [0.44–22.18] | 0.255 | 0.753 | 2.05 [0.25–16.72] | 0.501 | 0.999 | Yes |
| Staring | Autism without ID vs non-autism | 0.33 [0.04–2.46] | 0.277 | 0.753 | 0.37 [0.05–2.88] | 0.339 | 0.865 | Yes |
| Combativeness | Autism without ID vs non-autism | 3.14 [0.38–26.01] | 0.289 | 0.753 | 2.56 [0.29–22.59] | 0.398 | 0.963 | Yes |
| Stereotypy | Autism without ID vs non-autism | 2.80 [0.37–21.32] | 0.320 | 0.774 | 1.38 [0.14–13.93] | 0.784 | 0.999 | Yes |
| Mitgehen | Autism without ID vs non-autism | 3.24 [0.24–43.05] | 0.373 | 0.858 | 3.90 [0.27–56.08] | 0.316 | 0.865 | Yes |
| Withdrawal | Autism without ID vs non-autism | 0.44 [0.04–4.57] | 0.488 | 0.962 | 0.49 [0.04–5.29] | 0.554 | 0.999 | Yes |
| Mutism | Autism without ID vs non-autism | 1.94 [0.28–13.39] | 0.502 | 0.962 | 1.69 [0.23–12.11] | 0.604 | 0.999 | Yes |
| Negativism | Autism without ID vs non-autism | 1.80 [0.26–12.41] | 0.553 | 0.993 | 1.11 [0.14–8.90] | 0.922 | 0.999 | Yes |
| Waxy flexibility | Autism without ID vs non-autism | 1.85 [0.16–21.71] | 0.626 | 0.993 | 1.85 [0.15–22.99] | 0.633 | 0.999 | Yes |
| Excitement | Autism without ID vs non-autism | 1.64 [0.14–18.81] | 0.693 | 0.999 | 1.12 [0.09–14.17] | 0.929 | 0.999 | Yes |
| Autonomic abnormality | Autism without ID vs non-autism | 0.75 [0.07–7.91] | 0.809 | 0.999 | 1.25 [0.10–15.25] | 0.859 | 0.999 | No |
| Perseveration | Autism without ID vs non-autism | 1.26 [0.18–8.92] | 0.819 | 0.999 | 0.86 [0.11–6.84] | 0.890 | 0.999 | No |
| Posturing/catalepsy | Autism without ID vs non-autism | 1.17 [0.16–8.34] | 0.875 | 0.999 | 0.98 [0.13–7.31] | 0.984 | 0.999 | No |
| Rigidity | Autism without ID vs non-autism | 1.16 [0.15–8.71] | 0.888 | 0.999 | 1.15 [0.15–8.97] | 0.893 | 0.999 | Yes |
| Immobility/stupor | Autism without ID vs non-autism | 1.12 [0.16–7.84] | 0.908 | 0.999 | 1.32 [0.18–9.64] | 0.785 | 0.999 | Yes |
| Echopraxia/echolalia | Autism without ID vs non-autism | Not estimable | - | - | Not estimable | - | - | Not assessable |
| Grimacing | Autism without ID vs non-autism | Not estimable | - | - | Not estimable | - | - | Not assessable |
| Ambitendency | Autism without ID vs non-autism | Not estimable | - | - | Not estimable | - | - | Not assessable |
| Verbigeration | Autism without ID vs non-autism | Not estimable | - | - | Not estimable | - | - | Not assessable |
| Automatic obedience | Autism without ID vs non-autism | Not estimable | - | - | Not estimable | - | - | Not assessable |
| Gegenhalten | Autism without ID vs non-autism | Not estimable | - | - | Not estimable | - | - | Not assessable |
| Grasp reflex | Autism without ID vs non-autism | Not estimable | - | - | Not estimable | - | - | Not assessable |

| **Clinical group by consultation setting** | | | | |
| --- | --- | --- | --- | --- |
| **Clinical group** | **Inpatient n (%)** | **Outpatient n (%)** | **Total** | **Structural association** |
| Autism with ID | 8/36 (22.2%) | 28/36 (77.8%) | 36 | χ²=37.69, df=2, p=<0.001, Cramér's V=0.602 |
| Autism without ID | 2/5 (40.0%) | 3/5 (60.0%) | 5 |  |
| Non-autism | 53/63 (84.1%) | 10/63 (15.9%) | 63 |  |

**Flexible-age sensitivity for primary autism-with-intellectual-disability findings**

| **BFCRS item** | **N** | **Flexible-age adjusted OR [95% CI]** | **Raw p** | **FDR q** | **FDR significant** |
| --- | --- | --- | --- | --- | --- |
| **Stereotypy** | 104 | **14.03 [3.55–55.46]** | <0.001 | **0.004** | **Yes** |
| **Mannerisms** | 104 | **12.19 [3.28–45.29]** | <0.001 | **0.004** | **Yes** |
| **Impulsivity** | 104 | **8.91 [2.66–29.89]** | <0.001 | **0.006** | **Yes** |
| **Excitement** | 104 | **12.32 [2.97–51.10]** | <0.001 | **0.006** | **Yes** |
| **Perseveration** | 104 | **5.71 [1.79–18.27]** | 0.003 | **0.030** | **Yes** |
| **Negativism** | 104 | **5.70 [1.73–18.76]** | 0.004 | **0.032** | **Yes** |
| **Combativeness** | 104 | **7.44 [1.83–30.28]** | 0.005 | **0.033** | **Yes** |
| **Verbigeration** | 104 | **5.30 [1.58–17.74]** | 0.007 | **0.039** | **Yes** |
| Immobility/stupor | 104 | 0.21 [0.06–0.70] | 0.011 | 0.057 | No |

*Age at consultation was modeled using a restricted cubic spline with three degrees of freedom. Models otherwise retained the primary adjustment set. Benjamini-Hochberg FDR correction retained the original 46-contrast family. The panel includes the nine autism-with-intellectual-disability findings that survived FDR correction in the primary linear-age analysis.*

a *Primary adjusted models included three-level clinical group, age at consultation, biologic sex, and calendar year of electroconvulsive therapy initiation. Non-autism was the reference group.*

b *Structural-access sensitivity models additionally included inpatient versus outpatient consultation setting. These estimates were treated as sensitivity analyses because equivalent inpatient electroconvulsive therapy pathways were not readily available to autistic patients with intellectual disability.*

c *Benjamini-Hochberg false-discovery rate correction was applied independently within the 46 primary clinical-group contrasts and the 46 consultation-setting-adjusted clinical-group contrasts. Boldface identifies primary autism-with-intellectual-disability versus non-autism estimates that remained significant at q<0.05. No setting-adjusted contrast survived FDR correction.*

d *Direction was considered preserved when the primary and setting-adjusted log odds ratios had the same sign, corresponding to both odds ratios being either above 1 or below 1. Direction was preserved for 21 of 23 autism-with-intellectual-disability versus non-autism item estimates.*

e *Estimates for autism without intellectual disability should be interpreted as exploratory because this subgroup included five patients. Models affected by sparse-data separation are reported as not estimable rather than displaying unstable numerical estimates.*

***Abbreviations:*** *BFCRS, Bush-Francis Catatonia Rating Scale; CI, confidence interval; ECT, electroconvulsive therapy; FDR, false-discovery rate; ID, intellectual disability; OR, odds ratio.*
