## Supplemental Table 4 for "Catatonic Features That May Be Mistaken for Worsening Autism Show Treatment-Associated Improvement: Implications for Later Regression"

**Supplemental Table 4. BFCRS Longitudinal Psychomotor-Domain and Total-Score Analyses**

Values are regression coefficients β [95% CI]. Negative coefficients indicate decreasing scores with increasing ln(days+1). Interaction columns compare each autism subgroup with non-autism.

**Panel A. Unrestricted primary longitudinal models**

| **Outcome** | **Assessments** | **Patients** | **Non-autism slope** | **Autism with ID slope** | **Autism with ID vs non-autism interaction** | **Autism without ID slope** | **Autism without ID vs non-autism interaction** |
| --- | --- | --- | --- | --- | --- | --- | --- |
| Increased psychomotor behavior | 983 | 96 | -0.024 [-0.066, 0.018]; p=0.261; q=0.290 | -0.073 [-0.116, -0.030]; p=<0.001; q=0.001 | -0.049 [-0.109, 0.011]; p=0.109; q=0.598 | -0.139 [-0.287, 0.009]; p=0.067 | -0.115 [-0.269, 0.039]; p=0.145; q=0.598 |
| Abnormal psychomotor behavior | 985 | 96 | -0.055 [-0.086, -0.025]; p=<0.001; q=<0.001 | -0.072 [-0.098, -0.045]; p=<0.001; q=<0.001 | -0.016 [-0.056, 0.024]; p=0.427; q=0.598 | -0.062 [-0.126, 0.001]; p=0.055 | -0.007 [-0.078, 0.063]; p=0.842; q=0.842 |
| Decreased psychomotor behavior | 985 | 96 | -0.077 [-0.122, -0.031]; p=<0.001; q=0.001 | -0.060 [-0.080, -0.040]; p=<0.001; q=<0.001 | 0.017 [-0.033, 0.066]; p=0.515; q=0.598 | -0.124 [-0.270, 0.021]; p=0.093 | -0.048 [-0.200, 0.104]; p=0.538; q=0.598 |
| Autonomic abnormality | 984 | 96 | -0.018 [-0.051, 0.015]; p=0.293; q=0.293 | -0.040 [-0.080, 0.001]; p=0.056; q=0.070 | -0.022 [-0.074, 0.030]; p=0.411; q=0.598 | -0.045 [-0.119, 0.028]; p=0.225 | -0.028 [-0.108, 0.053]; p=0.505; q=0.598 |
| Total BFCRS | 985 | 96 | -1.269 [-2.004, -0.535]; p=<0.001; q=0.001 | -1.555 [-2.027, -1.084]; p=<0.001; q=<0.001 | -0.286 [-1.159, 0.587]; p=0.520; q=0.598 | -2.009 [-3.929, -0.089]; p=0.040 | -0.740 [-2.791, 1.311]; p=0.479; q=0.598 |

**Panel B. Unrestricted longitudinal models with additional adjustment for consultation setting**

| **Outcome** | **Assessments** | **Patients** | **Non-autism slope** | **Autism with ID slope** | **Autism with ID vs non-autism interaction** | **Autism without ID slope** | **Autism without ID vs non-autism interaction** |
| --- | --- | --- | --- | --- | --- | --- | --- |
| Increased psychomotor behavior | 983 | 96 | -0.025 [-0.069, 0.018]; p=0.254 | -0.074 [-0.117, -0.030]; p=<0.001 | -0.048 [-0.109, 0.013]; p=0.123; q=0.604 | -0.143 [-0.292, 0.006]; p=0.059 | -0.118 [-0.272, 0.037]; p=0.135; q=0.604 |
| Abnormal psychomotor behavior | 985 | 96 | -0.055 [-0.087, -0.024]; p=<0.001 | -0.072 [-0.098, -0.045]; p=<0.001 | -0.016 [-0.058, 0.025]; p=0.439; q=0.604 | -0.066 [-0.129, -0.004]; p=0.038 | -0.011 [-0.080, 0.058]; p=0.755; q=0.755 |
| Decreased psychomotor behavior | 985 | 96 | -0.077 [-0.123, -0.031]; p=<0.001 | -0.060 [-0.080, -0.040]; p=<0.001 | 0.017 [-0.033, 0.067]; p=0.504; q=0.604 | -0.126 [-0.272, 0.020]; p=0.091 | -0.048 [-0.201, 0.104]; p=0.533; q=0.604 |
| Autonomic abnormality | 984 | 96 | -0.015 [-0.047, 0.018]; p=0.369 | -0.039 [-0.079, 0.001]; p=0.057 | -0.024 [-0.075, 0.027]; p=0.350; q=0.604 | -0.041 [-0.111, 0.030]; p=0.260 | -0.026 [-0.104, 0.053]; p=0.522; q=0.604 |
| Total BFCRS | 985 | 96 | -1.278 [-2.039, -0.517]; p=<0.001 | -1.556 [-2.027, -1.084]; p=<0.001 | -0.278 [-1.175, 0.620]; p=0.544; q=0.604 | -2.073 [-3.977, -0.168]; p=0.033 | -0.795 [-2.828, 1.239]; p=0.444; q=0.604 |

**Panel C. Common-window longitudinal sensitivity models**

| **Window, days** | **Outcome** | **Assessments** | **Patients** | **Non-autism slope** | **Autism with ID slope** | **Autism with ID vs non-autism interaction** | **Autism without ID slope** | **Autism without ID vs non-autism interaction** |
| --- | --- | --- | --- | --- | --- | --- | --- | --- |
| 30 | Increased psychomotor behavior | 390 | 89 | -0.039 [-0.071, -0.008]; p=0.014; q=0.022 | -0.080 [-0.144, -0.015]; p=0.015; q=0.022 | -0.040 [-0.112, 0.031]; p=0.272; q=0.453 | -0.140 [-0.290, 0.010]; p=0.068 | -0.100 [-0.254, 0.053]; p=0.200; q=0.445 |
|  | Abnormal psychomotor behavior | 391 | 89 | -0.083 [-0.113, -0.054]; p=<0.001; q=<0.001 | -0.087 [-0.136, -0.038]; p=<0.001; q=<0.001 | -0.004 [-0.061, 0.054]; p=0.900; q=0.900 | -0.093 [-0.128, -0.058]; p=<0.001 | -0.009 [-0.055, 0.037]; p=0.692; q=0.769 |
|  | Decreased psychomotor behavior | 391 | 89 | -0.135 [-0.194, -0.076]; p=<0.001; q=<0.001 | -0.078 [-0.149, -0.007]; p=0.031; q=0.039 | 0.057 [-0.035, 0.149]; p=0.222; q=0.445 | -0.215 [-0.283, -0.147]; p=<0.001 | -0.080 [-0.170, 0.010]; p=0.082; q=0.412 |
|  | Autonomic abnormality | 390 | 89 | -0.042 [-0.101, 0.018]; p=0.169; q=0.188 | 0.029 [-0.042, 0.100]; p=0.420; q=0.420 | 0.071 [-0.022, 0.163]; p=0.133; q=0.443 | -0.068 [-0.182, 0.046]; p=0.240 | -0.027 [-0.155, 0.102]; p=0.686; q=0.769 |
|  | Total BFCRS | 391 | 89 | -2.056 [-2.670, -1.442]; p=<0.001; q=<0.001 | -1.818 [-2.778, -0.858]; p=<0.001; q=<0.001 | 0.238 [-0.903, 1.379]; p=0.683; q=0.769 | -2.986 [-3.310, -2.663]; p=<0.001 | -0.930 [-1.625, -0.236]; p=0.009; q=0.086 |
| 60 | Increased psychomotor behavior | 494 | 91 | -0.047 [-0.090, -0.004]; p=0.034; q=0.042 | -0.081 [-0.134, -0.028]; p=0.003; q=0.004 | -0.035 [-0.103, 0.034]; p=0.319; q=0.532 | -0.175 [-0.341, -0.009]; p=0.039 | -0.128 [-0.300, 0.043]; p=0.143; q=0.429 |
|  | Abnormal psychomotor behavior | 495 | 91 | -0.080 [-0.111, -0.049]; p=<0.001; q=<0.001 | -0.079 [-0.118, -0.040]; p=<0.001; q=<0.001 | 0.001 [-0.048, 0.051]; p=0.958; q=0.958 | -0.097 [-0.153, -0.040]; p=<0.001 | -0.016 [-0.081, 0.048]; p=0.618; q=0.772 |
|  | Decreased psychomotor behavior | 495 | 91 | -0.140 [-0.193, -0.086]; p=<0.001; q=<0.001 | -0.083 [-0.123, -0.044]; p=<0.001; q=<0.001 | 0.057 [-0.010, 0.123]; p=0.094; q=0.429 | -0.210 [-0.318, -0.102]; p=<0.001 | -0.071 [-0.191, 0.050]; p=0.251; q=0.501 |
|  | Autonomic abnormality | 494 | 91 | -0.040 [-0.090, 0.010]; p=0.116; q=0.129 | 0.016 [-0.031, 0.063]; p=0.505; q=0.505 | 0.056 [-0.013, 0.125]; p=0.110; q=0.429 | -0.043 [-0.125, 0.038]; p=0.298 | -0.003 [-0.099, 0.093]; p=0.947; q=0.958 |
|  | Total BFCRS | 495 | 91 | -2.078 [-2.736, -1.419]; p=<0.001; q=<0.001 | -1.753 [-2.454, -1.052]; p=<0.001; q=<0.001 | 0.325 [-0.637, 1.286]; p=0.508; q=0.726 | -3.095 [-4.396, -1.794]; p=<0.001 | -1.017 [-2.476, 0.441]; p=0.172; q=0.429 |
| 90 | Increased psychomotor behavior | 572 | 93 | -0.048 [-0.089, -0.007]; p=0.023; q=0.028 | -0.091 [-0.139, -0.043]; p=<0.001; q=<0.001 | -0.043 [-0.106, 0.021]; p=0.186; q=0.373 | -0.188 [-0.356, -0.019]; p=0.029 | -0.140 [-0.313, 0.033]; p=0.114; q=0.373 |
|  | Abnormal psychomotor behavior | 573 | 93 | -0.081 [-0.112, -0.049]; p=<0.001; q=<0.001 | -0.080 [-0.112, -0.048]; p=<0.001; q=<0.001 | 0.001 [-0.044, 0.045]; p=0.977; q=0.989 | -0.095 [-0.148, -0.043]; p=<0.001 | -0.015 [-0.076, 0.047]; p=0.641; q=0.801 |
|  | Decreased psychomotor behavior | 573 | 93 | -0.138 [-0.192, -0.084]; p=<0.001; q=<0.001 | -0.082 [-0.115, -0.050]; p=<0.001; q=<0.001 | 0.056 [-0.007, 0.119]; p=0.082; q=0.373 | -0.207 [-0.307, -0.106]; p=<0.001 | -0.068 [-0.182, 0.046]; p=0.240; q=0.400 |
|  | Autonomic abnormality | 572 | 93 | -0.041 [-0.082, 0.001]; p=0.055; q=0.061 | 0.007 [-0.025, 0.040]; p=0.655; q=0.655 | 0.048 [-0.005, 0.101]; p=0.076; q=0.373 | -0.041 [-0.119, 0.037]; p=0.303 | -0.001 [-0.089, 0.088]; p=0.989; q=0.989 |
|  | Total BFCRS | 573 | 93 | -2.076 [-2.756, -1.396]; p=<0.001; q=<0.001 | -1.794 [-2.380, -1.207]; p=<0.001; q=<0.001 | 0.282 [-0.616, 1.181]; p=0.538; q=0.768 | -3.091 [-4.303, -1.880]; p=<0.001 | -1.016 [-2.406, 0.375]; p=0.152; q=0.373 |
| 180 | Increased psychomotor behavior | 719 | 95 | -0.054 [-0.093, -0.014]; p=0.008; q=0.009 | -0.103 [-0.152, -0.053]; p=<0.001; q=<0.001 | -0.049 [-0.112, 0.014]; p=0.127; q=0.423 | -0.179 [-0.334, -0.024]; p=0.024 | -0.125 [-0.285, 0.035]; p=0.125; q=0.423 |
|  | Abnormal psychomotor behavior | 721 | 95 | -0.083 [-0.111, -0.055]; p=<0.001; q=<0.001 | -0.077 [-0.109, -0.045]; p=<0.001; q=<0.001 | 0.006 [-0.036, 0.048]; p=0.784; q=0.957 | -0.078 [-0.142, -0.015]; p=0.016 | 0.005 [-0.065, 0.074]; p=0.894; q=0.957 |
|  | Decreased psychomotor behavior | 721 | 95 | -0.122 [-0.170, -0.073]; p=<0.001; q=<0.001 | -0.073 [-0.100, -0.047]; p=<0.001; q=<0.001 | 0.048 [-0.007, 0.103]; p=0.087; q=0.423 | -0.162 [-0.298, -0.026]; p=0.019 | -0.041 [-0.185, 0.104]; p=0.581; q=0.831 |
|  | Autonomic abnormality | 720 | 95 | -0.039 [-0.071, -0.007]; p=0.018; q=0.020 | -0.022 [-0.050, 0.006]; p=0.126; q=0.126 | 0.017 [-0.026, 0.059]; p=0.439; q=0.831 | -0.041 [-0.115, 0.033]; p=0.279 | -0.002 [-0.083, 0.079]; p=0.957; q=0.957 |
|  | Total BFCRS | 721 | 95 | -2.027 [-2.626, -1.428]; p=<0.001; q=<0.001 | -1.761 [-2.353, -1.169]; p=<0.001; q=<0.001 | 0.266 [-0.575, 1.107]; p=0.536; q=0.831 | -2.569 [-4.297, -0.842]; p=0.004 | -0.542 [-2.370, 1.286]; p=0.561; q=0.831 |

**Panel D. Complete-assessment longitudinal sensitivity models**

| **Outcome** | **Assessments** | **Patients** | **Non-autism slope** | **Autism with ID slope** | **Autism with ID vs non-autism interaction** | **Autism without ID slope** | **Autism without ID vs non-autism interaction** |
| --- | --- | --- | --- | --- | --- | --- | --- |
| Increased psychomotor behavior | 891 | 94 | -0.019 [-0.064, 0.025]; p=0.394; q=0.394 | -0.067 [-0.109, -0.025]; p=0.002; q=0.004 | -0.048 [-0.109, 0.013]; p=0.127; q=0.660 | -0.138 [-0.285, 0.010]; p=0.067 | -0.118 [-0.272, 0.036]; p=0.132; q=0.660 |
| Abnormal psychomotor behavior | 891 | 94 | -0.049 [-0.082, -0.017]; p=0.003; q=0.005 | -0.066 [-0.093, -0.040]; p<0.001; q<0.001 | -0.017 [-0.059, 0.025]; p=0.434; q=0.695 | -0.062 [-0.126, 0.002]; p=0.058 | -0.012 [-0.084, 0.059]; p=0.733; q=0.733 |
| Decreased psychomotor behavior | 891 | 94 | -0.078 [-0.126, -0.030]; p=0.002; q=0.004 | -0.050 [-0.073, -0.028]; p<0.001; q<0.001 | 0.028 [-0.026, 0.081]; p=0.308; q=0.695 | -0.124 [-0.271, 0.022]; p=0.096 | -0.046 [-0.200, 0.108]; p=0.556; q=0.695 |
| Autonomic abnormality | 891 | 94 | -0.017 [-0.054, 0.020]; p=0.361; q=0.394 | -0.033 [-0.072, 0.005]; p=0.086; q=0.108 | -0.016 [-0.068, 0.036]; p=0.536; q=0.695 | -0.050 [-0.129, 0.029]; p=0.215 | -0.033 [-0.120, 0.054]; p=0.461; q=0.695 |
| Total BFCRS | 891 | 94 | -1.174 [-1.966, -0.382]; p=0.004; q=0.005 | -1.393 [-1.879, -0.907]; p<0.001; q<0.001 | -0.219 [-1.148, 0.711]; p=0.645; q=0.716 | -1.993 [-3.912, -0.075]; p=0.042 | -0.819 [-2.888, 1.250]; p=0.438; q=0.695 |

**Panel E. Independence working-correlation longitudinal sensitivity models**

| **Outcome** | **Assessments** | **Patients** | **Non-autism slope** | **Autism with ID slope** | **Autism with ID vs non-autism interaction** | **Autism without ID slope** | **Autism without ID vs non-autism interaction** |
| --- | --- | --- | --- | --- | --- | --- | --- |
| Increased psychomotor behavior | 983 | 96 | 0.008 [-0.041, 0.057]; p=0.758; q=0.758 | -0.076 [-0.122, -0.030]; p=0.001; q=0.003 | -0.084 [-0.149, -0.019]; p=0.011; q=0.115 | -0.119 [-0.253, 0.015]; p=0.082 | -0.127 [-0.266, 0.013]; p=0.076; q=0.337 |
| Abnormal psychomotor behavior | 985 | 96 | -0.029 [-0.065, 0.007]; p=0.119; q=0.149 | -0.067 [-0.095, -0.038]; p<0.001; q<0.001 | -0.038 [-0.083, 0.007]; p=0.101; q=0.337 | -0.063 [-0.128, 0.002]; p=0.056 | -0.034 [-0.107, 0.038]; p=0.353; q=0.504 |
| Decreased psychomotor behavior | 985 | 96 | -0.054 [-0.119, 0.012]; p=0.109; q=0.149 | -0.042 [-0.066, -0.019]; p<0.001; q=0.001 | 0.011 [-0.059, 0.081]; p=0.755; q=0.838 | -0.131 [-0.270, 0.008]; p=0.064 | -0.078 [-0.231, 0.076]; p=0.321; q=0.504 |
| Autonomic abnormality | 984 | 96 | -0.033 [-0.073, 0.007]; p=0.107; q=0.149 | -0.038 [-0.072, -0.004]; p=0.029; q=0.057 | -0.005 [-0.055, 0.045]; p=0.844; q=0.844 | -0.069 [-0.171, 0.033]; p=0.187 | -0.036 [-0.145, 0.074]; p=0.525; q=0.656 |
| Total BFCRS | 985 | 96 | -0.705 [-1.684, 0.273]; p=0.158; q=0.175 | -1.391 [-1.882, -0.899]; p<0.001; q<0.001 | -0.685 [-1.767, 0.396]; p=0.214; q=0.429 | -2.036 [-3.712, -0.359]; p=0.017 | -1.330 [-3.233, 0.573]; p=0.171; q=0.427 |

***a*** *Gaussian generalized estimating equation models used an identity link, patient-level clustering, and robust standard errors. Panels A–D used an exchangeable working-correlation structure; Panel E used an independence working-correlation structure.*

***b*** *Models included ln(days+1), three-level clinical group, the clinical-group-by-time interaction, age at consultation, biologic sex, and calendar year of electroconvulsive therapy initiation. Non-autism was the reference group.*

***c*** *Panel B additionally adjusted for inpatient versus outpatient consultation setting. Panel B group-specific slope p-values are nominal.*

***d*** *Group-specific slope p-values were adjusted within separate 10-test principal families for the unrestricted primary analysis, complete-assessment sensitivity analysis, independence working-correlation sensitivity analysis, and each 30-, 60-, 90-, and 180-day common follow-up window. Each family comprised five outcomes in the autism-with-intellectual-disability and non-autism groups. Autism-without-intellectual-disability slopes were exploratory and were not included in these families.*

***e*** *Clinical-group-by-time interaction p-values were adjusted within separate 10-test families for the unrestricted primary analysis, consultation-setting sensitivity analysis, complete-assessment sensitivity analysis, independence working-correlation sensitivity analysis, and each common follow-up window. Each family comprised five outcomes and two autism-versus-non-autism contrasts. No interaction survived false-discovery rate correction at q<0.05.*

***f*** *Coefficients represent change per one-unit increase in ln(days+1) and should not be interpreted as change per calendar day.*

***g*** *The increased domain included excitement, impulsivity, and combativeness. The abnormal domain included posturing/catalepsy, grimacing, echopraxia/echolalia, stereotypy, mannerisms, verbigeration, automatic obedience, mitgehen, gegenhalten, grasp reflex, perseveration, rigidity, and waxy flexibility. The decreased domain included immobility/stupor, mutism, staring, negativism, withdrawal, and ambitendency. Autonomic abnormality was analyzed separately.*

***h*** *Multi-item domain scores were calculated as mean item scores when at least 80% of constituent items were observed. Total BFCRS scores were prorated to 23 items for assessments with at least 20 observed items. Panel D was restricted to assessments with all 23 BFCRS items observed and therefore used no score proration.*

***i*** *Across all 104 patients, 993 near-complete BFCRS assessments were available. Unrestricted analyses were limited to 96 patients with at least two eligible assessments, leaving 985 assessments; outcome-specific unrestricted models included 983 to 985 assessments because of item-level missingness. Complete-assessment analyses included 891 assessments from 94 patients.*

***Abbreviations:*** *BFCRS, Bush-Francis Catatonia Rating Scale; CI, confidence interval; ECT, electroconvulsive therapy; FDR, false-discovery rate; GEE, generalized estimating equation; ID, intellectual disability.*
